# Copy number signature analyses in prostate cancer reveal distinct etiologies and clinical outcomes

**DOI:** 10.1101/2020.04.27.20082404

**Authors:** Shixiang Wang, Huimin Li, Minfang Song, Zaoke He, Tao Wu, Xuan Wang, Ziyu Tao, Kai Wu, Xue-Song Liu

## Abstract

Genome alteration signatures reflect recurring patterns caused by distinct endogenous or exogenous mutational events during the evolution of cancer. Signatures of single base substitution (SBS) have been extensively studied in different types of cancer, however, signatures of cancer genome copy number alteration (CNA) are still elusive in most cancer types, especially in prostate cancer (PC), which is particularly driven by complex genome alterations. Here, a user-friendly open source bioinformatics tool “sigminer” has been constructed for copy number signature extraction, analysis and visualization. Five copy number signatures are identified from human PC genome with this tool. The underlying driving forces for each signature have been illustrated. Sample clustering based on copy number signature exposure revealed considerable heterogeneity of PC, and copy number signatures show improved PC clinical outcome association when compared with SBS signatures. Copy number signature analyses provide distinct insight into the etiology of PC, and potential biomarkers for PC stratification and prognosis.

## Introduction

Prostate cancer (PC) is a common cancer type among men (Torre et al., 2015). Although some of these cancers are slow-growing, others are more aggressive and PC is annually responsible for a quarter million deaths world-wide (Jemal et al., 2011). One of the biggest unmet clinical needs in PC is to stratify clinically indolent from aggressive types, particularly in patients diagnosed at young age. Molecular markers have shown promise in risk stratification, but the utility is complicated by the heterogeneous natural history of PC, which develops over decades(Pound et al., 1999). Initial patient evaluation and treatment decisions are currently based on a risk stratification scheme that incorporates three prognostic biomarkers at diagnosis: clinical stage, biopsy Gleason score, and serum PSA(Gaudreau et al., 2016). However, these conventional biomarkers could not provide a precise risk stratification for PC patients.

Comprehensive genomic characterizations provide biomarkers for PC classification or stratification (Abida et al., 2019; Cancer Genome Atlas Research, 2015; Robinson et al., 2015; Taylor et al., 2010). Abeshouse *et al*. classified prostate tumors into 7 subtypes by ETS fusions or mutations in SPOP, FOXA1, and IDH1, and still a significant fraction of PC patients are driven by as-yet-unexplained molecular alterations. In addition, clinical outcomes associated with these 7 PC subsets are not clear(Cancer Genome Atlas Research, 2015).

Copy number alteration (CNA) is an important driver for the progression of multiple types of cancer (Beroukhim et al., 2010; Zack et al., 2013). PCs also have varying degrees of DNA copy number alteration; indolent and low-Gleason tumors have few alterations, whereas more aggressive primary and metastatic tumors have extensive copy number alterations (Hieronymus et al., 2014; Taylor et al., 2010). In contrast, somatic point mutations are less common in PC than in most other solid tumors (Grasso et al., 2012). The most frequently mutated genes in primary PC are *SPOP, TP53, FOXA1*, and *PTEN* (Barbieri et al., 2012).

Recently, several studies reported that percent of copy number altered genome, termed “CNA burden” is associated with the recurrence and death of prostate tumors (Hieronymus et al., 2014) and other tumor types (Hieronymus et al., 2018). Aneuploidy, defined as chromosome gains and losses, has been reported to drive lethal progression in prostate cancer (Stopsack et al., 2019). All these results suggest a key role of CNA in prostate cancer progression and clinical outcome association. However, the underlying mechanisms remain unknown. It is also unclear whether there are some specific types of copy number alterations that are more likely to be associated with the clinical outcomes of PC.

To explore and address these challenges, we developed a novel method to investigate the signatures of copy number alterations, and build an open source R/CRAN package for the scientific community. Our method incorporated the copy number features developed by previous studies (Korbel and Campbell, 2013; Macintyre et al., 2018; Menghi et al., 2016), and also included new features to reflect the chromosomal distribution pattern of copy number alterations. In addition, a copy number feature component counting based method has been applied in signature extraction, and this facilitates not only computational efficiency and signature visualization, but also integration with other signature analysis tools including SBS signature analysis. The driving forces and mechanisms underlying each distinct copy number signature have been proposed, the clinical outcome for each signature has been further investigated. This copy number signature analysis provides a new insight into the etiology of prostate tumors, and also novel biomarkers for PC stratification and prognosis.

## Results

### Copy number signature analysis framework

The distinct events that drive copy number alterations in human cancers are not readily identifiable from high-throughput sequencing data. This presents a major challenge for the development of precision medicine for cancers that are strongly affected by copy number alterations, including prostate cancer. Recent studies have enabled the interpretation of genomic changes by identifying mutational signatures— genomic patterns that are the imprint of mutagenic processes accumulated over the lifetime of a cancer cell (Alexandrov et al., 2020; Alexandrov et al., 2013).

SBS mutations are caused by single-strand DNA lesions and single-strand break repair mistakes. Copy number alterations reflect double-strand DNA lesions and double-strand break repair problems. SBS signatures have been well studied in many cancers, and summarized in the COSMIC database (Alexandrov et al., 2020). However, copy number signatures are less studied except a recent report performed copy number signature analysis in high-grade serous ovarian cancer (Macintyre et al., 2018).

Here a novel copy number signature extraction method has been constructed (Figure 1). It incorporated the following 8 copy number features: the breakpoint count per 10 Mb (named “BP10MB”); the breakpoint count per chromosome arm (named “BPArm”); the absolute copy number of the segments (named “CN”); the difference in copy number values between adjacent segments (named copy number change point, or “CNCP”); the lengths of oscillating copy number segment chains (named “OsCN”); the log10 based size of segments (named “SS”); the minimal number of chromosomes with 50% copy number alterations (named “NC50”); the distribution of copy number alterations in each chromosome (burden of chromosome, named “BoChr”) (Figure 1). These features were selected as hallmarks of previously reported genomic aberrations like chromothripsis, tandem duplication or to denote the genome distribution pattern of copy number alteration events (Korbel and Campbell, 2013; Macintyre et al., 2018; Menghi et al., 2016).

**Figure 1.**
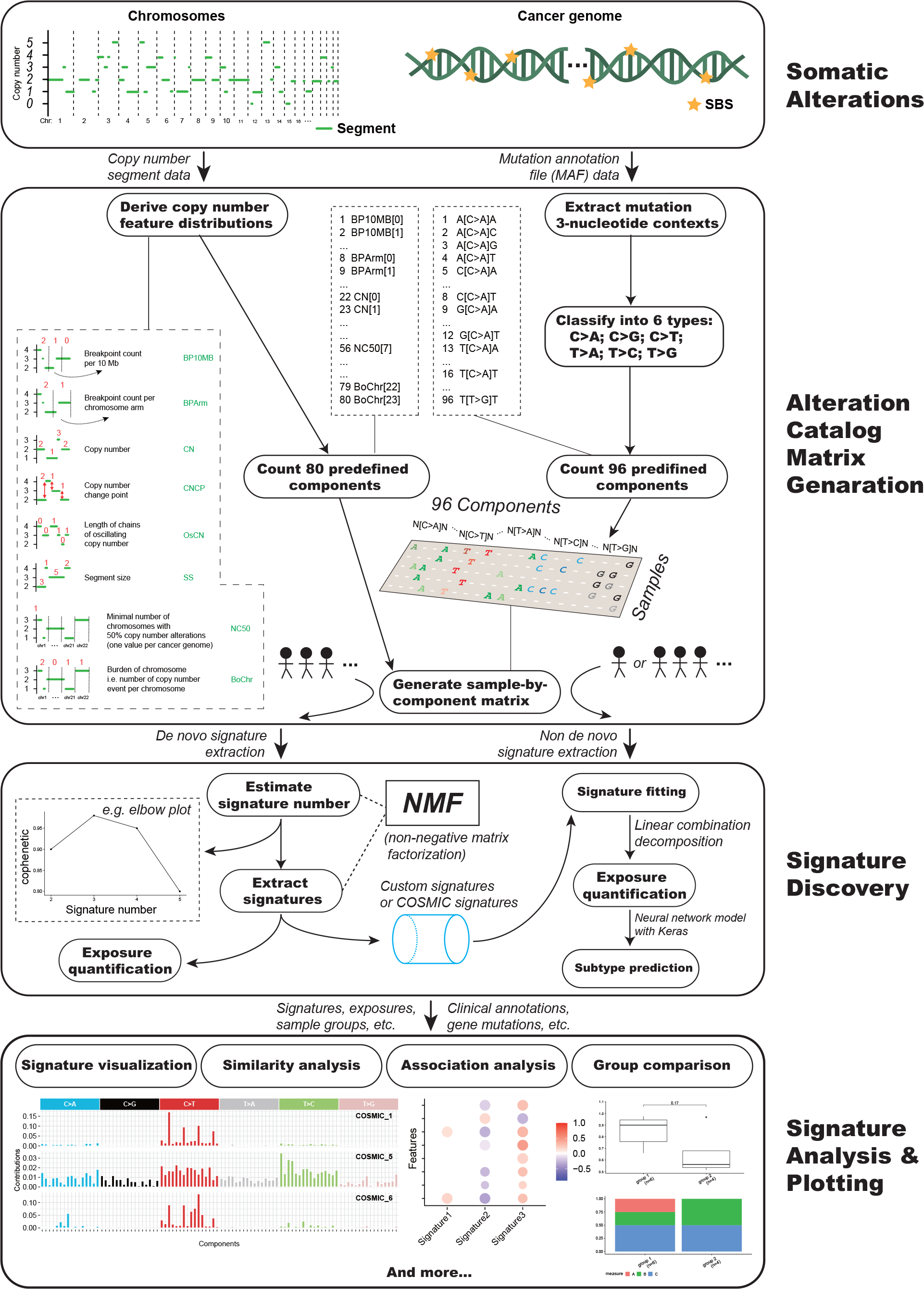
Genome alteration signature analysis framework.

We classified the distributions of the above mentioned 8 copy number features into 80 components after comprehensive analysis of the value range, abundance and biological significance of each copy number feature (Figure 1 and Table S1). For each patient, the value for each component of copy number feature was counted based on the segmented absolute copy number profile of that patient. The absolute copy number profiles can be extracted from whole exome sequencing (WES), whole genome sequencing (WGS), or SNP array data (Favero et al., 2015; Shen and Seshan, 2016).Then a sample by copy number component value matrix was generated by combining component values in all samples. This matrix was subjected to non-negative matrix factorization (NMF), a method previously used for deriving SBS signatures (Alexandrov et al., 2013). SBS signatures were derived in a similar way from a counting matrix including 96 components.

Compared to previous copy number signature extraction method, our method is much more computationally efficient, it took about 1.5 minutes with our method rather than about 1.5 hours using Macintyre et al method to generate the matrix for copy number signature extraction. Besides, each of the predefined 80 copy number components has clear biological meaning, and this facilitates not only the intelligibility of signature result, but also the scalability of this copy number signature analysis method with known SBS signature analysis tool. A user-friendly R/CRAN package (sigminer) has been developed based on this novel copy number analysis method for bioinformatics community and cancer researchers to explore and analyze both SBS signatures and copy number signatures (Figure 1). This tool is released under MIT license, and all source code is freely available on GitHub. To our knowledge, sigminer is the first practical bioinformatics tool for extracting the signatures of copy number alterations.

### Genome alteration landscape of PC

We assembled and uniformly analyzed WES data from 1,003 pairs of prostate cancers and matched germline control (649 primary and 354 metastatic tumors) that passed quality control parameters from six independent studies (Methods, Figure S1) (Beltran et al., 2011; Berger et al., 2011; Cancer Genome Atlas Research, 2015; Grasso et al., 2012; Kohli et al., 2015; Robinson et al., 2015). Patient characteristics, including age at diagnosis, Gleason score, and metastatic site, are shown in Table S2.

Small scale genome alterations include single base substitution (SBS) and small insertion and deletion (INDEL). Mutsig analysis with these small scale genome alterations revealed 47 cancer driver genes (MutSig q < 0.05) (Figure S2). Similar to previous study(Barbieri et al., 2012), the top frequently mutated genes in this PC genome dataset are *TP53* (19%), *SPOP* (9%), *FOXA1* (8%), *PTEN* (4%) (Figure S2). About half of PC samples (56.44%, 552 of 978) have small scale cancer driving genome alterations. These small scale genome alterations were summarized based on different variant classifications (Figure S3A-C). The median number of small scale variants in PC is 28, and most of them are SBS, suggesting a relatively low SBS number in PC compared with other cancer types (Figure S3B).

Distribution pattern of CNA segment length and chromosome distribution of CNA counts in available PC samples are summarized and shown (Figure S3D). Majority of copy number alterations are focal, and 14.9% are chromosome arm level or whole chromosome level alterations (Figure S3D). Median values of CNA segment count, CNA amplification count, CNA deletion count and CNA burden are shown in Table S3. The distribution of 8 CNA features selected in this study are shown (Figure S3E).

### Genome alteration signatures identified in PC

With the integrated copy number signature and SBS signature analysis tool sigminer developed in this study, we identified five copy number signatures and three SBS signatures from 1003 tumor-normal pairs of PC WES data. The number of signature was determined after comprehensive consideration of result stability shown by cophenetic plot and biological interpretability (Figure S4).

Three SBS mutational signatures named SBS-sig 1, SBS-sig 2, SBS-sig 3 are identified (Figure 2). Etiology for the SBS mutational signatures has been well explored and stored in COSMIC database. Cosine similarity analysis has been performed between the three SBS signatures identified in this study and COSMIC signatures (Figure S5). SBS-Sig 1 is similar to COSMIC signature 3 or 5. COSMIC signature 3 correlates to defects in DNA repair by homologous recombination and the etiology of COSMIC signature 5 is unknown. SBS-Sig 2 is similar to COSMIC signature 15 or 6, and both of them are related to defective DNA mismatch repair. SBS-Sig 3 is similar to COSMIC signature 1, which is featured by spontaneous and age-related deamination of 5-methylcytosine. According to signature matrix provided by COSMIC, three COSMIC signatures, i.e. COSMIC signature 1, 5, and 6 were identified according to TCGA prostate cancer data. Thus, the SBS signatures identified in this study are similar to those identified by COSMIC based on TCGA PC datasets.

**Figure 2.**
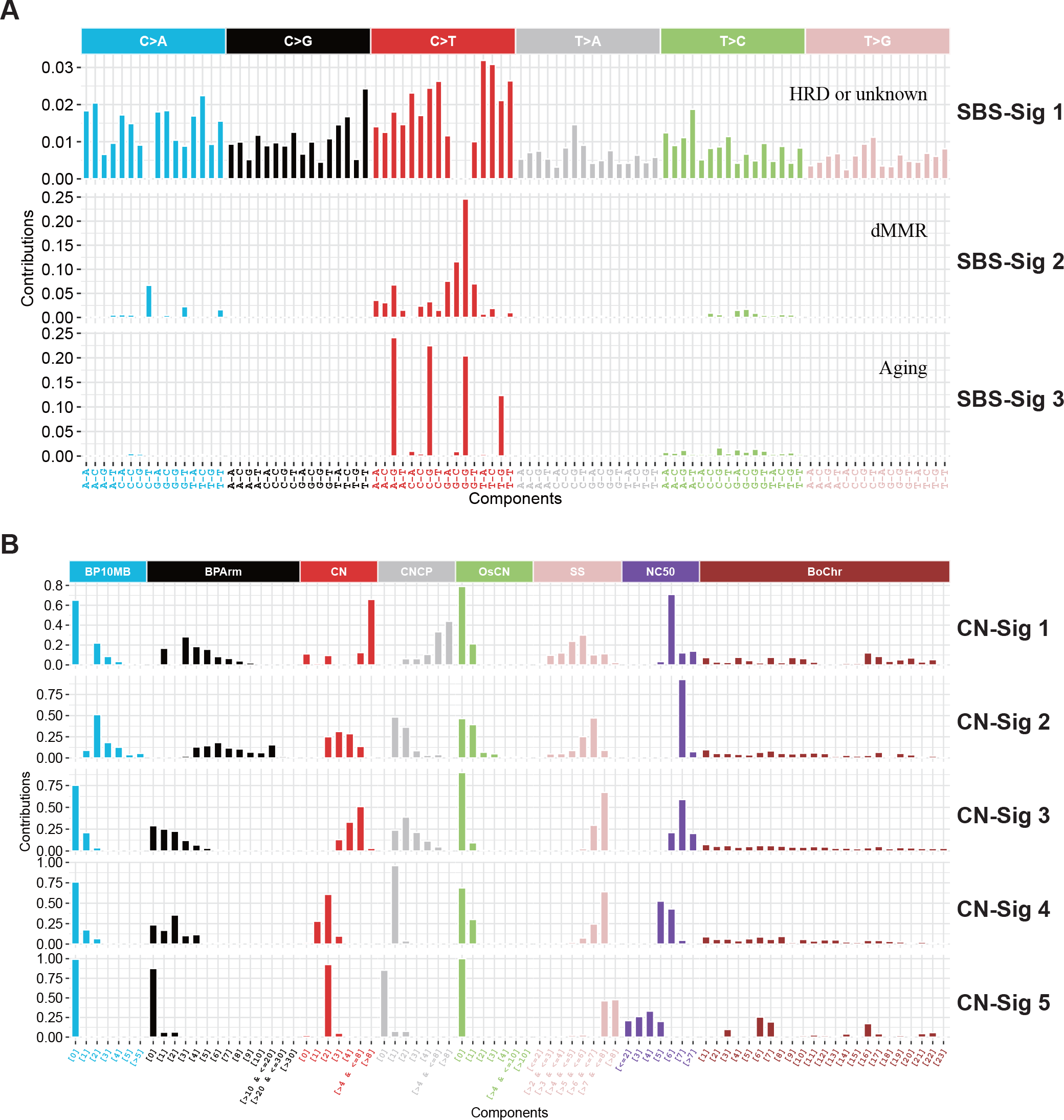
SBS signatures and copy number signatures identified in PC. (A, B) Three SBS signatures (A) and five copy number signatures (B) are identified from prostate cancer WES data. For SBS signatures, the data was row normalized; for copy number signatures, the data was row normalized within each feature. All 96 components in SBS signature belong to one feature, while copy number signature has 8 different features with 80 components in together.

Five copy number alteration signatures are identified, namely CN-sig 1 to CN-sig 5 (Figure 2). These copy number signatures are ranked based on the median length of CNA segment. CN-sig 1 has the smallest median size of CNA segment, and CB-sig 5 has the largest median size of CNA segment. Copy number components of the same feature are row normalized, and this facilitates copy number component value comparisons within the signature. Macintyre *et al*. performed column normalization for each copy number component, and the component values of same CNA feature cannot be compared within each signature (Macintyre et al., 2018). Representative absolute copy number profile for each signature enriched PC patient is shown (Figure S6). The copy number signatures identified in this study can be validated with an independent PC dataset which has segmented copy number profiles but not raw WES sequencing data available for analysis (MSKCC2020, http://www.cbioportal.org/study/summary?id=prad_mskcc_2020) (Figure S7). We found highly similar component weights for the signatures in this independent PC cohort, and this demonstrates the robustness of both the methodology and the copy number features (Figure S8A and B, median similarity = 0.90), despite a significant difference in exposures to copy number signatures between the cohorts (P < 0.05, two-sided Mann-Whiney U test, Figure S8C).

### Mutational processes underlying copy number alteration signatures

Five copy number signatures and three SBS mutational signatures have been identified in 1003 tumor-normal paired PC WES dataset. These signatures reflect distinct genomic DNA alteration patterns driving by distinct molecular events. To investigate the interconnection among genome alteration signatures, we computed the associations between the exposures of signatures and various types of genomic alteration features including tumor purity, MATH (A simple quantitative measure of intra-tumor heterogeneity (Mroz and Rocco, 2013)), tumor ploidy, CNA burden, INDEL number, tandem duplication phenotype score (TDP score, see methods) (Menghi et al., 2016), chromothripsis state score (see methods) (Korbel and Campbell, 2013) (Figure 3A), genetic alteration in known PC driving genes (Figure 3B), and PC associated signaling pathways (Figure 3C and Table S4).

**Figure 3.**
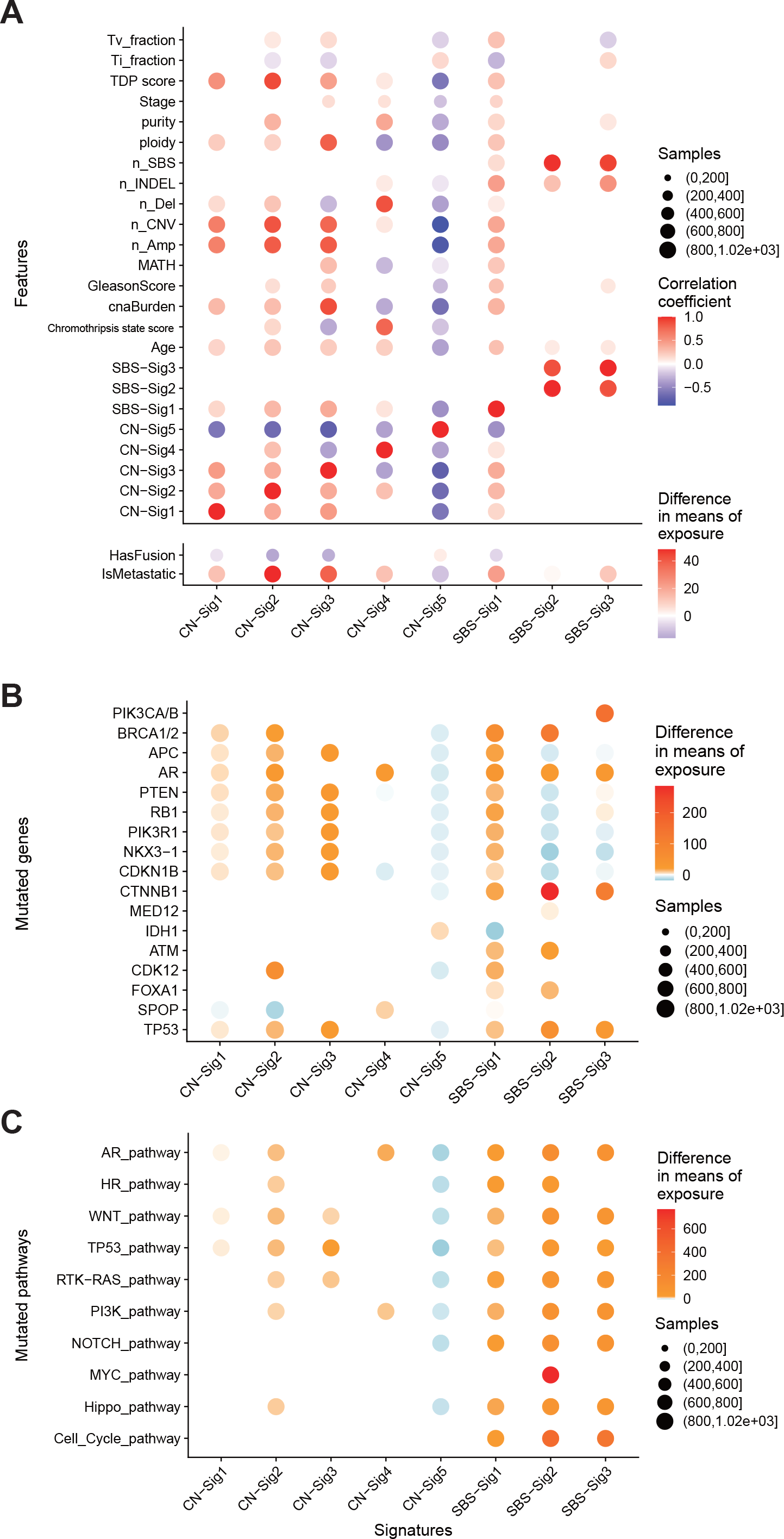
Genome alteration signatures and mutational processes in PC. (A) Associations between the exposures of genome alteration signatures and features including clinical parameters, somatic DNA alteration quantifications. Only Pearson correlations with false-discovery rate P < 0.05 are shown. (B, C) Associations between the exposures of genome alteration signatures and mutations in specific genes (B) or selected pathways (C). Only differences with false-discovery rate P < 0.05 (Mann–Whitney U-test) are shown. Red, positive correlation; blue, negative correlation. The depths of colors in filled circles indicate the extent of difference. Sizes of circle in all plots indicate the number of cases included in each analysis.

CN-sig 5 show negative association with nearly all cancer genome alteration features excluding *TMPRSS2-ETS* oncogenic fusions. And CN-sig 5 is the only genome alteration signatures that show positive correlation with the presence of *TMPRSS2-ETS* oncogenic fusions. *TMPRSS2-ETS* fusions happens in early stage of PC (Gerhauser et al., 2018). All these observations suggest that CN-sig 5 reflecting a stable genome or early PC evolving state. CN-sig 4 show strongest association with number of genome deletions. CN-sig 3 show strongest positive correlation with tumor ploidy and CNA burden. CN-sig 2 show strongest association with PC metastasis.

Multidimensional clustering with genome alteration signatures and PC clinical parameters results in several clusters (Figure S9). CNA burden feature is clustered with CN-sig 1, 2, 3, 5. And in this cluster, CN-sig 5 show negative correlations with other features or signatures. CN-sig 4 is associated with number of deletions, and form a distinct cluster when compared to other copy number signatures. SBS signatures form separate clusters from copy number signatures. SBS-sig 2 and SBS-sig 3 clustered with number of INDELs, total mutation, and SBS-sig 1 form a different cluster. These analyses revealed underlying connections between different genome alteration signatures.

Based on the distribution of the values of each copy number components (Figure 2), copy number profiles of representative samples (Figure S6), and signature correlation analyses (Figure 4 and Figure S9), the etiology and mechanism for CN signature has been proposed as following (Table 1):

**Table 1.**
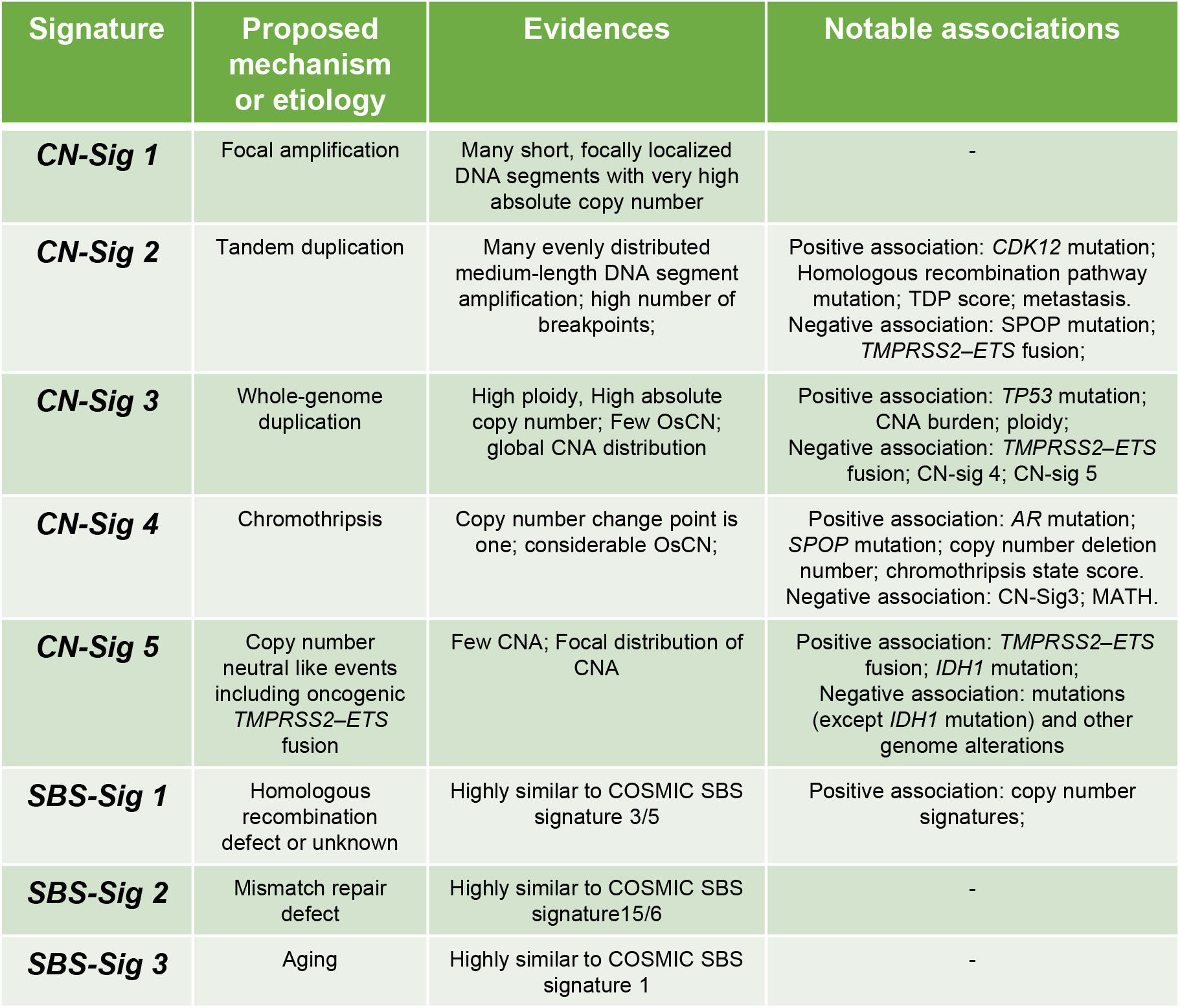
Genome alteration signatures in prostate cancer. Associations for each genome alteration signature and proposed mechanisms are shown.

| Signature | Proposed mechanism or etiology | Evidences | Notable associations |
| --- | --- | --- | --- |
| **CN-Sig 1** | Focal amplification | Many short, focally localized DNA segments with very high absolute copy number | - |
| **CN-Sig 2** | Tandem duplication | Many evenly distributed medium-length DNA segment amplification; high number of breakpoints; | Positive association: *CDK12* mutation; Homologous recombination pathway mutation; TDP score; metastasis. Negative association: *SPOP* mutation; *TMPRSS2-ETS* fusion; |
| **CN-Sig 3** | Whole-genome duplication | High ploidy, High absolute copy number; Few OsCN; global CNA distribution | Positive association: *TP53* mutation; CNA burden; ploidy; Negative association: *TMPRSS2-ETS* fusion; CN-sig 4; CN-sig 5 |
| **CN-Sig 4** | Chromothripsis | Copy number change point is one; considerable OsCN; | Positive association: *AR* mutation; *SPOP* mutation; copy number deletion number; chromothripsis state score. Negative association: CN-Sig3; MATH. |
| **CN-Sig 5** | Copy number neutral like events including oncogenic *TMPRSS2-ETS* fusion | Few CNA; Focal distribution of CNA | Positive association: *TMPRSS2-ETS* fusion; *IDH1* mutation; Negative association: mutations (except *IDH1* mutation) and other genome alterations |
| **SBS-Sig 1** | Homologous recombination defect or unknown | Highly similar to COSMIC SBS signature 3/5 | Positive association: copy number signatures; |
| **SBS-Sig 2** | Mismatch repair defect | Highly similar to COSMIC SBS signature15/6 | - |
| **SBS-Sig 3** | Aging | Highly similar to COSMIC SBS signature 1 | - |

**Figure 4.**
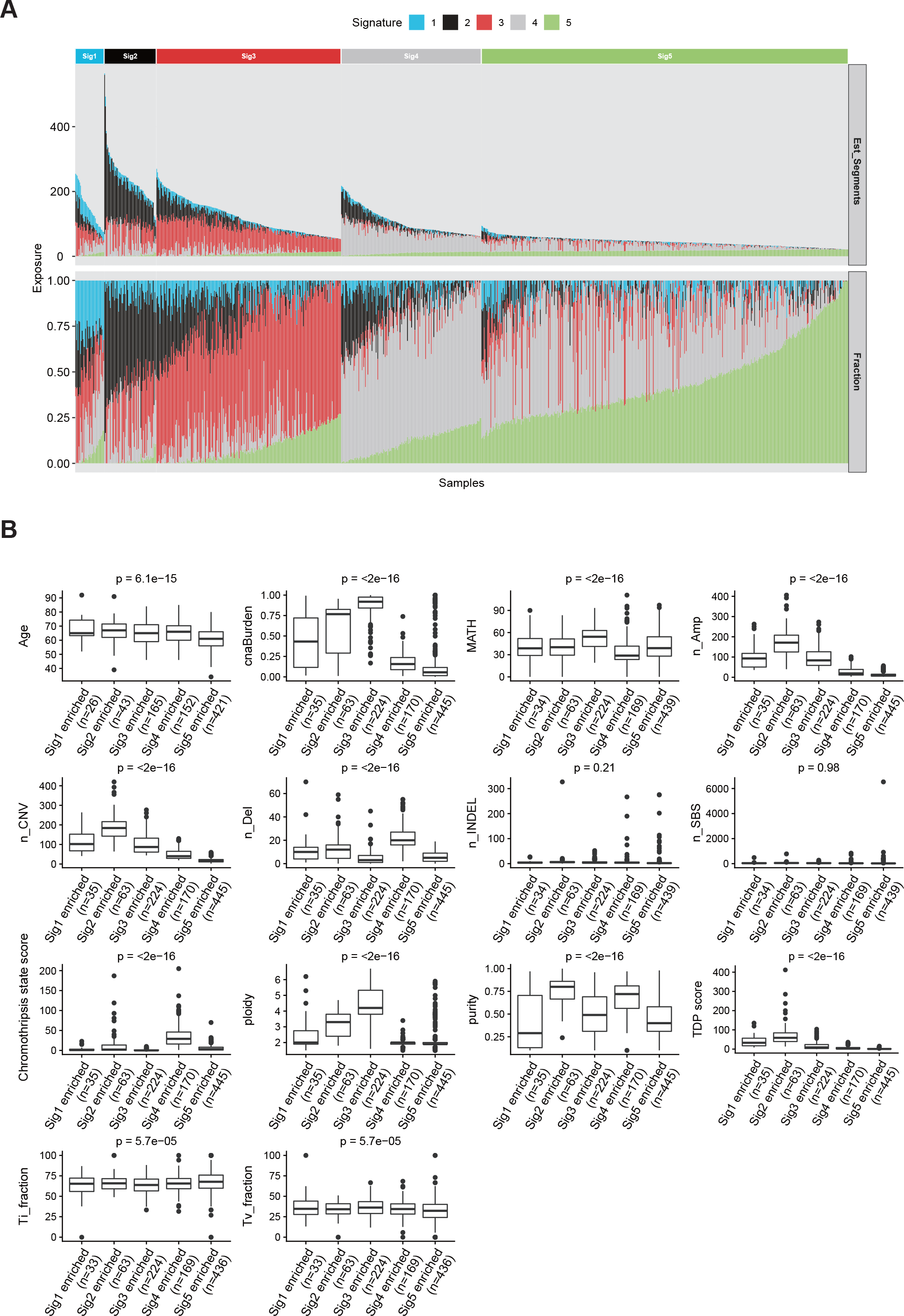
Sample clustering and heterogeneity analysis of PC based on the exposures of copy number signatures. (A) For each PC patient, the relative contribution (bottom panel) and estimated copy number segment counts (top panel) of each signature are shown as a staked barplot. PC samples are clustered into five groups based on the consensus matrix from multiple NMF runs, and each group is specified by one enriched copy number signature. (B) Quantification comparison for somatic and clinical parameters among each copy number signature enriched PC group by boxplot. ANOVA p values are shown. Abbr.: n_, number of, e.g. n_SNV, number of SNV; INDELs, small insertions and deletions; Ti_fraction, transition fraction; Tv_fraction, transversion fraction; Amp, copy number segments with amplification; Del, copy number segments with deletion; TDP score, tandem duplication phenotype score; cnaBurden, copy number alteration burden; MATH, MATH score is a quantitative measure of intra-tumor heterogeneity.

CN-sig 1 is represented by many short (0.01-0.1Mb), focally localized DNA segments with very high absolute copy number (>8). This signature is caused by focal amplification of DNA segments, and some of them can be caused by the formation of extrachromosomal DNA (ecDNA), and this need further investigation (Deshpande et al., 2019; Turner et al., 2017).

CN-sig 2 is represented by evenly or globally distributed, large amount of medium-length (0.1-10 Mb) DNA segment amplifications. This signature is associated with *CDK12*, homologous recombination (HR) pathway gene mutations and high TDP score, and show highest association with the metastasis of PC. This signature can be a result of defective HR DNA repair and consequently tandem duplication phenotypes.

CN-sig 3 is represented by high tumor ploidy, and is associated with high CNA burden and *TP53* mutation. This signature reflects the occurrence of whole genome doubling events.

CN-sig 4 is featured by DNA segment one copy deletion, and considerable oscillating copy number. This signature is associated with *AR, SPOP* mutation and high chromothripsis state score, suggesting a state of chromothripsis (Yi and Ju, 2018).

CN-sig 5 is represented by large copy number segment size with few background copy number alterations, and is the only signature positively associated with PC oncogenic fusions. This signature is also the only copy number signature enriched specifically in non-metastatic PC, and is associated with good survival. This signature is a result of relatively stable PC genomes, reflecting copy number neutral mutation processes like *TMPRSS2-ETS* oncogenic fusions.

### Copy number signature and PC patients’ stratification

PC samples are clustered into five groups based on the consensus matrix from multiple NMF runs, and each group is specified by one enriched copy number signature (Figure 4A). Clinical and genomic parameters including patients’ age, SBS count (n_SBS), number of insertion/deletions (n_INDEL), transition (Ti) fraction, transversion (Tv) fraction, CNA number, copy number amplification number (n_Amp), copy number deletion number (n_Del), CNA burden, tumor purity, and tumor ploidy are compared in each copy number signature enriched PC patient group (Figure 4B). Significant differences are observed in all above mentioned clinical parameters, except total mutation and number of INDEL among different copy number signature enriched PC group (Figure 4B). Similar analysis was performed when clustering PC patients with SBS signatures. Most clinical and genomic parameters do not show significant difference among three SBS signature enriched PC groups (Figure S10). This analysis suggests that copy number signatures could have more PC stratification power when compared with SBS signatures.

Metastasis is the leading cause of PC associated death, the associations between each genome alteration signatures and PC metastasis are shown as Sankey diagram (Figure 5A). CN-Sig 2 are nearly exclusively found to be associated with PC metastasis, and CNA-Sig 5 are nearly exclusively found not to be associated with PC metastasis. SBS mutational signatures do not show strong associations with PC metastasis as copy number signatures. This metastasis association analysis suggested that copy number signature can be PC metastasis specific biomarker, and this analysis further demonstrates that copy number signatures provide more PC stratification information than SBS mutational signature.

**Figure 5.**
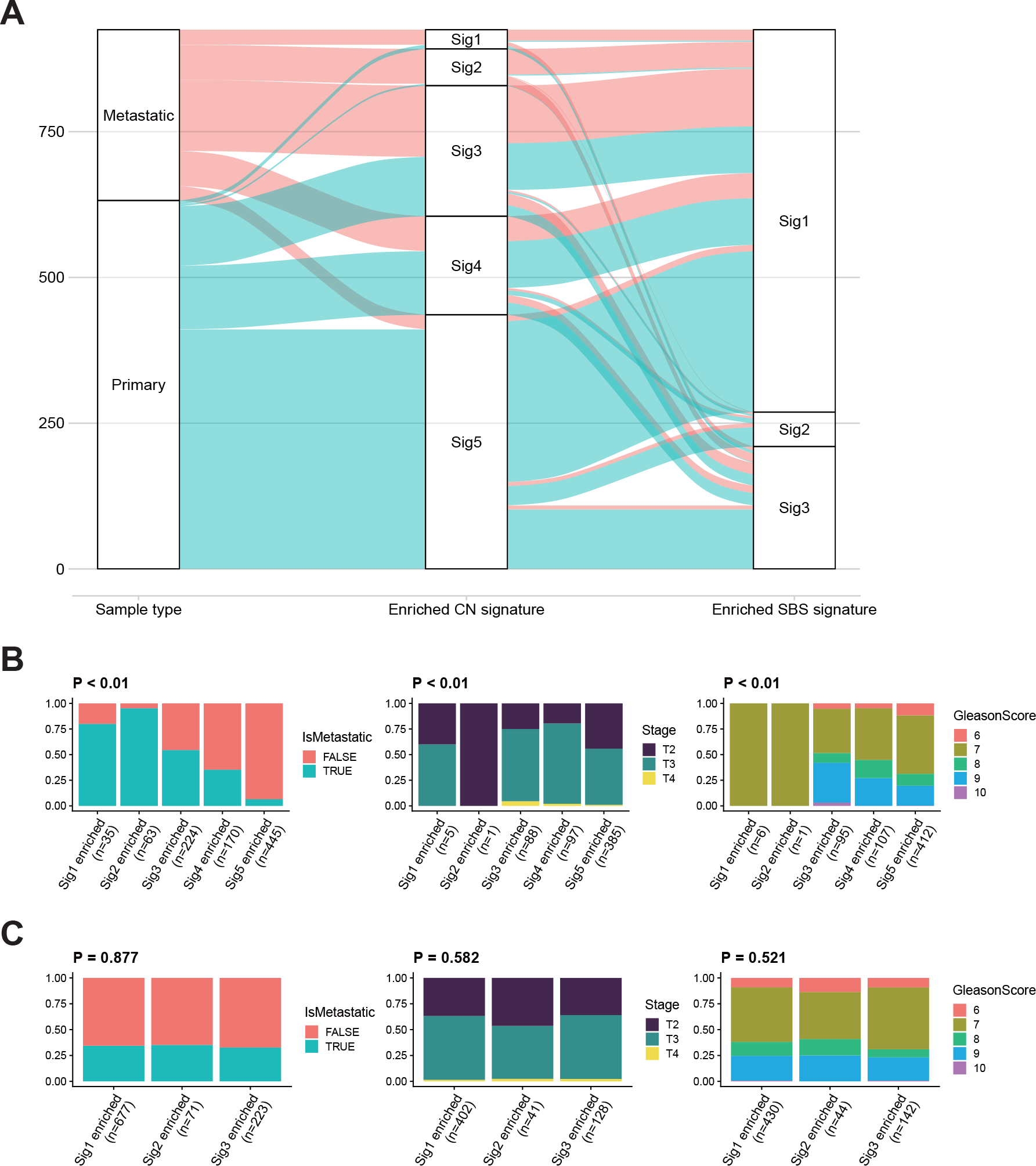
Associations between genome alteration signatures and PC clinical variables including metastatic status, clinical stage and Gleason score. (A) Relationship between copy number signature exposure or SBS signature exposure and metastasis is shown by a Sankey plot. The sample size are indicated on the left of plot. (B, C) Fraction changes of clinical variables including metastasis, clinical stage and Gleason score in each copy number signature enriched PC groups (B) or each SBS signature enriched PC groups (C). p values were calculated by Fisher test with Monte Carlo simulation.

The associations between copy number signatures and metastasis, Gleason score and clinical stage have been statistically analyzed (Figure 5B). Similar analysis has been performed with SBS mutational signatures (Figure 5C). Significant differences in metastasis, Gleason score and clinical stage status are observed among different copy number signature enriched PC patients. However, among SBS mutational signature enriched PC patients, no significant differences are observed. These analyses again demonstrate that copy number signatures carry more PC stratification information than SBS mutational signatures.

### Copy number signatures predict PC patients’ survival

Univariate Cox regression analyses were performed to evaluate the associations between each genome alteration signature and PC patients’ survival time (Figure 6). In overall survival (OS) analysis, only copy number signatures show significant correlations, none of SBS signatures show significant correlations (Figure 6A). CN-sig 2 is highly significantly (P<0.001) (hazard ratio=1.14, 95% CI 1.08-1.20) associated with poor OS, and CN-sig 5 is highly significantly (P<0.001) (hazard ratio=0.88, 95% CI 0.83-0.93) associated with improved OS. In regards to progression-free survival (PFS), CN-sig 3 and CN-sig 5 show significant correlations, CN-sig 3 is associated with poor PFS (hazard ratio=1.09, 95% CI 1.04-1.14, P<0.001), and CN-sig 5 is associated with improved PFS (hazard ratio=0.92, 95% CI 0.88-0.97, P=0.002) (Figure 6B). CN-sig 2 tends to be associated with poor PFS, and this association is not statistically significant based on the datasets used in this study (hazard ratio=1.29, 95% CI 0.94-1.76, P=0.116). SBS-sig 1, SBS-sig 2 are significantly associated with poor PFS (Figure 6B).

**Figure 6.**
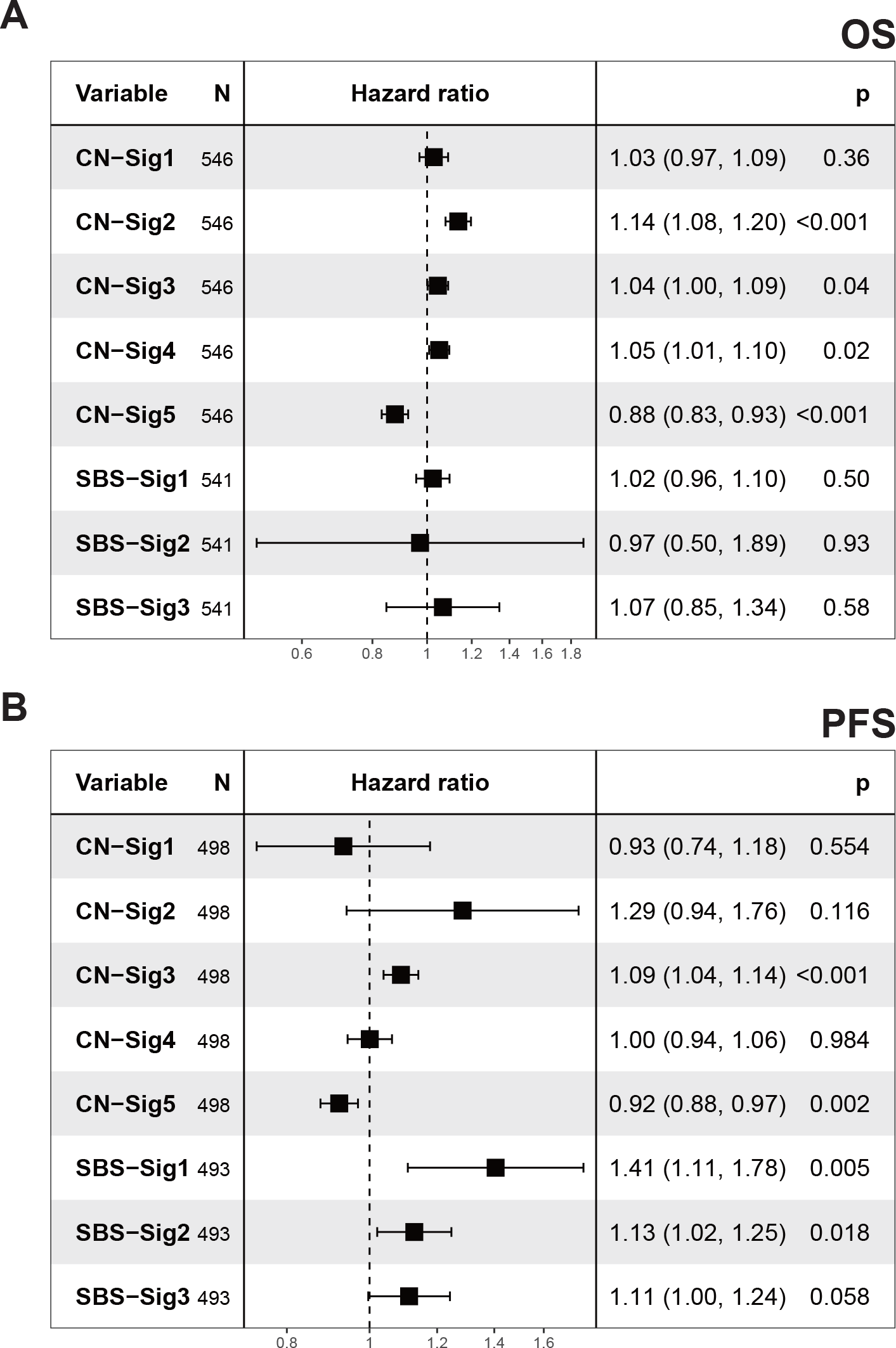
Genome alteration signature exposures and PC survival. (A, B) Forest plots showing the relative risk of signature exposures in overall survival (OS) (A) and progression-free survival (PFS) (B). Exposures for all signatures were normalized to 1-20 for evaluating the Hazard ratios per 5% exposure increase. Hazard ratios and p values were calculated by univariable Cox analysis.

The associations between other clinical genomic parameters and OS, PFS are analyzed with univariate Cox regression method (Figure S11). As expected, high Gleason scores are significantly associated with both poor OS and poor PFS. Late clinical stages are significantly associated with poor PFS. CNA burden is significantly associated with poor PFS. High tumor purity is significantly associated with poor OS, and high tumor ploidy is significantly associated with poor PFS.

This survival time analysis suggests that copy number signatures are associated with PC patients’ survival. CN-sig 5 is associated with improved survival, while CN sig-2 is associated with poor survival. This is consistent with previous studies which shows that increased CNA burden is associated with poor PC survival (Hieronymus et al., 2014). In addition, this study further suggest that specific type of CNA alteration represented by CN-sig 2 is associated with poor PC overall survival. CN-sig 2 has the highest hazard ratio among all five copy number signatures in both OS and PFS analysis. Of note, CN-sig 2 is not the copy number signature with the highest CNA burden. CN-sig 3 has the highest CNA burden, as expected CN-sig 3 is associated with poor survival, however the hazard ratio for CN-sig 3 is lower than the hazard ratio for CN-sig 2 in both OS and PFS analysis (Figure 6).

## Discussion

Here a novel copy number alteration signature analysis method has been constructed. This method is featured by enhanced signature extraction efficiency, enhanced component visualization and comprehension, and can be coherently integrated with the known SBS signature analysis method. With this newly developed bioinformatics tool “sigminer”, we performed the first copy number signature analysis in PC, and identified five copy number signatures. Copy number signatures show improved performance in PC patients’ stratification and survival prediction compared with SBS signatures. Importantly, copy number signatures can be specific biomarkers for PC metastasis. These analyses provide novel insight into the etiology of PC, and novel biomarkers for PC stratification and prognosis. We demonstrated the utility of “sigminer” in PC, and this tool can be applied in other cancer types for the deep understanding of the complex genomic DNA alterations.

Genomic DNA alteration signatures are recurring genomic patterns that are the imprint of mutagenic processes accumulated over the lifetime of a cancer cell. The underlying driving forces for each signature have been proposed based on previous evidence and association study. The driving forces for SBS mutational signature have been well-studied, cosine similarity analysis indicates that the three SBS signatures identified in this study are similar to those reported in COSMIC database based on TCGA data. The copy number signatures associated with chromothripsis, tandem duplication, and focal amplification have been identified in PC. The proposed mechanisms for each identified copy number signature need more experimental and clinical evidence to further validate.

In OS analysis, the prognostic performance of CNA signatures is much improved compared with SBS signatures. In addition, copy number signatures also show improved association with PC metastasis and clinical stages compared with SBS signature. In together, copy number signatures show improved PC clinical outcome association when compared with SBS signature. And this difference needs to be further investigated in other cancer types. This implicates that the factors underlying copy number alterations can be major driving forces for PC progression and metastasis.

Our copy number signature analysis tool “sigminer” only needs absolute copy number profile derived from WES or SNP array data, and high coverage WGS data is not needed. However, the detailed structural alteration information derived from recent pan cancer WGS project can help the future improvement of copy number signature analysis (Li et al., 2020). Signature analysis for other types of genomic DNA alterations, such as insertion/deletion, SV and their integration with SBS and CNA signature need further study. The correlation between genome alteration signatures and patients’ precision medicine need continuous investigation. DNA alteration signature can ultimately improve cancer patients’ stratification, clinical outcome prognosis, therapy outcome prediction.

## Materials and Methods

### Cohort collection and preprocessing

PC samples were included in this study if tumor and matched germline whole exome sequencing (WES) raw data (BAM or SRA files) were accessible. In total, 6 cohorts are included in this study, their dbGaP accession numbers are phs000178 (TCGA) (Cancer Genome Atlas Research, 2015), phs000447 (Berger et al., 2011), phs000554 (Grasso et al., 2012), phs000909 (Beltran et al., 2011), phs000915 (Robinson et al., 2015) and phs001141 (Kohli et al., 2015). In total 1003 tumor-normal paired WES data are available for this study. BAM files for TCGA cohort were downloaded from GDC portal (https://portal.gdc.cancer.gov/) with tool gdc-client. The average sequencing depth of each BAM files are summarized in Table S5. SRA files for other cohorts were obtained from dbGaP database (https://dbgap.ncbi.nlm.nih.gov/) and converted to FASTQ files by SRA toolkit. Adapters were removed from the FASTQ files by trimgalore (https://github.com/FelixKrueger/TrimGalore). BWA MEM algorithm were then applied using hg38 as reference genome (Li, 2013). The result SAM files were converted to BAM files with samtools (Li et al., 2009), followed by Picard toolkit (https://broadinstitute.github.io/picard/) to sort BAM files and mark duplications for variant and copy number calling. Details for processing steps and the corresponding codes are provided in section “Code availability”. All tumor samples were required to have at least 50X mean target coverage, and all paired normal samples were required to have at least 30X mean target coverage.

### Clinical data

Clinicopathological annotations for cohorts phs000447, phs000554, phs000909, phs000915 and phs001141 were obtained from the original papers and dbGap database. Clinicopathological annotations for TCGA prostate cohort were downloaded from UCSC Xena by R package UCSCXenaTools v1.2.10 (Wang and Liu, 2019). The data were cleaned and organized by in-house R scripts available in “Code availability” section.

### Absolute copy number calling from WES

Two bioinformatics tools including Sequenza (Favero et al., 2015) and FACETS (Shen and Seshan, 2016) were performed to generate absolute copy number profile from tumor-normal paired WES BAM files. For Sequenza, we followed its standard pipeline described in its vignette (https://cran.r-project.org/web/packages/sequenza/vignettes/sequenza.html) but with parameter ‘female’ set to ‘FALSE’ and a modified R package copynumber (https://github.com/ShixiangWang/copynumber) to work with hg38 genome build. For FACETS, we strictly followed its standard workflow described in its vignette (https://github.com/mskcc/facets/blob/master/vignettes/FACETS.pdf). Samples failed in calling were excluded from downstream analysis. In total Sequenza successfully called 937 pairs of WES data, and FACETS called 933 pairs of WES data. Finally, the absolute copy number data generated by Sequenza was selected for downstream analysis in this study based on the fact that FACETS generated some unreasonable copy number profile for a few samples. The somatic absolute copy number profiles detected in this study are summarized in Table S6.

### Variant calling from WES

Somatic mutations including SBS (single base substitutions) and INDEL (insertions and deletions) were detected by following the best practices of Genome Analysis Toolkit (GATK v4.1.3) with Mutect2 (Benjamin et al., 2019). The lists of genome variants that passed all quality control filters were converted into VCF files by VCFtools (Danecek et al., 2011). The resulting VCF files were annotated by VEP (McLaren et al., 2016) and further converted to MAF file by vcf2maf.pl (https://github.com/mskcc/vcf2maf). The MAF file was loaded into R, analyzed and visualized by Maftools (Mayakonda et al., 2018). The somatic genome variants detected in this study are summarized in Table S7.

### Genome alteration signature extraction procedures

To identify both SBS and copy number signatures, a unified signature extraction framework has been developed and was implemented as an R/CRAN package sigminer (https://github.com/ShixiangWang/sigminer), which also contains features about signature analysis and visualization. Basically, *de novo* signature discovery can be divided into the following three steps.

1. Tally variation components. a) for SBS profile, same as previously reported (Alexandrov et al., 2020; Alexandrov et al., 2013), for each patient, we firstly classified mutation records into six substitution subtypes: C>A, C>G, C>T, T>A, T>C, and T>G (all substitutions are referred to by the pyrimidine of the mutated Watson–Crick base pair). Further, each of the substitutions was examined by incorporating information on the bases immediately 5’ and 3’ to each mutated base generating 96 possible mutation types (6 types of substitution ∗ 4 types of 5’ base ∗ 4 types of 3’ base). Each of 96 mutation types is called component here. b) For copy number profile, we firstly computed the genome-wide distributions of 8 fundamental copy number features for each patient: the breakpoint count per 10 Mb (named BP10MB); the breakpoint count per chromosome arm (named BPArm); the copy number of the segments (named CN); the difference in copy number between adjacent segments (named CNCP); the lengths of oscillating copy number segment chains (named OsCN); the log10 based size of segments (named SS); the minimal number of chromosome with 50% copy number variation (named NC50); the burden of chromosome (named BoChr). These features were selected as hallmarks of previously reported genomic aberrations like chromothripsis or to denote the distribution pattern of copy number events (Korbel and Campbell, 2013; Menghi et al., 2016; Murnane, 2012; Ng et al., 2012). The former 6 features have been used in previous study (Macintyre et al., 2018) to uncover the mutational processes in ovarian carcinoma. Next, unlike previous study (Macintyre et al., 2018), which applied mixture modeling to separate the first 6 copy number features distributions into mixtures of Poisson or Gaussian distributions, we directly classified 8 copy number features distributions into different components according to the comprehensive consideration of value range, abundance and biological significance. Most of the result are discrete values, and the remaining are range values (see Table S1). Based on genome alteration components defined above, two patient-by-component matrices (one for SBS and the other for copy number) were generated and treated as the input of non-negative matrix factorization (NMF) algorithm for extracting signatures as previously reported (Alexandrov et al., 2020; Alexandrov et al., 2013; Gaujoux and Seoighe, 2010).
2. Estimate signature number. Signature number (or factorization rank) is a critical value which affects both the performance of NMF algorithm and biological interpretability. A common way for deciding on signature number is to try different values, compute some quality measure of the results, and choose the best value according to this quality criteria (Gaujoux and Seoighe, 2010). The most common approach is to use the cophenetic correlation coefficient (Brunet et al., 2004). As suggested, performing 30-50 runs is considered sufficient to obtain a robust estimation of the signature number value (Gaujoux and Seoighe, 2010). We performed 50 runs for both SBS (signature number range from 2 to 10) and copy number signatures (signature number range from 2 to 12). According to the cophenetic vs signature number plot, the number of SBS signatures and copy number signatures were selected to be three and five respectively, based on both the indication of stability shown by cophenetic plot and biological interpretability.
3. Extract signatures. After determining the signature number, we performed NMF with 50 runs to extract signatures for downstream analysis.

### Signature profile normalization

Signature profile is essentially a matrix with row representing signature components and column representing the contributions of each signature component. Same as previous reported (Alexandrov et al., 2020), SBS signature profile was normalized within each signature (i.e. by row). However, it is harder to read and understand if the same method was applied to copy number signature profile due to the relative independence of each copy number feature. We performed row normalization within each copy number feature, so a component value can be compared to another component value of the same copy number feature within each signature. In summary, we did row normalization for SBS signature profile and row normalization within each feature for copy number signature, and generated a bar plot whose bar height indicates the observed component frequency compared to other components of the same feature.

### Signature exposure quantification and cancer subtype classification

Sigminer package can provide both relative and absolute exposures of each signature for cancer patient. The relative exposure represents the relative contribution of a signature in a sample. The absolute exposure estimates the information records a signature generated, for SBS signature, it is the SBS count; for copy number signature, it is the number of copy number segment. After *de novo* signature discovery, these two types of exposure information can be readily obtained with sigminer package. Sometimes, it would be useful to quantify the signature exposures for a new cancer sample based on existing signatures (for example from *de novo* signature discovery). sigminer package adopted the linear combination decomposition (LCD) method from Bioconductor package YAPSA to fit the identified signatures to one or more samples. To classify a new cancer sample based on signature exposures, we built a 5-layer neural network model (input layer + hidden layer + 2 dropout layers + output layer) with Keras library (https://keras.rstudio.com/) and trained it with datasets used in our study. This model can predict the subtype for single new prostate cancer sample with copy number data available with high accuracy. The steps to build and train the model for practical use are packaged and available at https://github.com/ShixiangWang/sigminer.prediction for research community.

### Association analysis

Same as previously reported (Macintyre et al., 2018), associations between the exposures of signatures and other clinical or genomic features was performed using one of two procedures: 1) for a continuous association variable (including ordinal variable like clinical stage), Pearson correlation was performed; 2) for a binary variable, patients were divided into two groups and a Mann-Whiney U-test was performed to test for differences in average exposures of signatures between the two groups.

### Correlation network analysis

To investigate the structure of signature associations, correlation network analysis was performed with R package corrr (https://github.com/tidymodels/corrr) with continuous association variables. Variables that are more correlated appear closer together and are joined by thicker curves. Red and blue curves indicate positive and negative correlation respectively. The proximity of the points was determined with multidimensional clustering.

### Score definitions

To quantify the tandem duplication (TD) status for each sample, we defined a tandem duplication phenotype (TDP) score, which capture the distribution and length information of TD across chromosomes (Menghi et al., 2018; Menghi et al., 2016). This TDP score is calculated as:

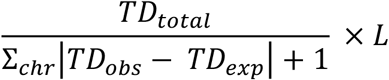

Where *TD*_*total*_ is the total number of TD in a sample, *TD*_*obs*_ and *TD*_*exp*_ are the observed and expected TD for a chromosome in the sample, *L* is the total size of TD in Mb unit. TD is defined as copy number amplification segments with size range from 1Kb to 2Mb.

To quantify the chromothripsis state for each sample, we defined a chromothripsis state score. This score is based on one key feature of chromothripsis, which forms tens to hundreds of locally clustered segmental losses being interspersed with regions displaying normal (disomic) copy-number (Korbel and Campbell, 2013). This chromothripsis state score is calculated as:

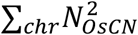

Where 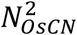 is the square of total number of copy number fragments with absolute copy number value following “2-1-2” pattern. This score is a simplified estimation of chromothripsis state for each sample.

### Copy number signature validation

To validate the copy number signatures identified in this study with independent dataset, we downloaded MSKCC 2020 PC cohort data from cBioPortal (https://www.cbioportal.org/study/summary?id=prad_cdk12_mskcc_2020). Segmented copy number profiles but not raw WES sequencing data are available, and this segmented copy number data for 1465 PC samples were used as the input of ABSOLUTE software to call absolute copy number profile for each PC sample (Carter et al., 2012). 163 PC samples failed to report absolute copy number profiles were filtered out for further analysis. As previously described, five copy number signatures were extracted. These five copy number signatures in MSKCC 2020 cohort were then compared with CN-Sig1 to CN-Sig5 with cosine similarity analysis. The exposures of each copy number signature are compared between MSKCC 2020 cohort and the collected datasets of this study.

### Survival analysis

Univariate Cox analysis and visualization were performed by R package survival and ezcox (https://github.com/ShixiangWang/ezcox). The cox model returns hazard ratio value (including 95% confidence interval) per variable unit increase. To properly determine the relevance between variables and PC patients’ OS and PFS, exposures of all signatures were normalized to 1-20 for indicating the HR per 5% exposure increase. In a similar way, we multiplied some variables including CNA burden, Ti fraction and tumor purity by 20 (the raw values of these variables range from 0 to 1). For other variables, we directly evaluated the HR per variable unit increase, e.g. HR for n_SBS indicates the HR per SBS count increase in a sample.

### Statistical analysis

Correlation analysis was performed using the Pearson method. Mann-Whiney U-test was performed to test for differences in signature exposure medians between the two groups. ANOVA was performed to test for differences across more than 2 groups. Fisher test was performed to test the association between signatures and categorical variables. In larger than 2 by 2 tables, Fisher p-values were calculated by Monte Carlo simulation. For multiple hypothesis testing, p values were adjusted using the FDR method. All reported p-values are two-tailed, and for all analyses, p<=0.05 is considered statistically significant, unless otherwise specified. Statistical analyses were performed by R v3.6 (https://cran.r-project.org/).

## Data Availability

All code required to reproduce the analysis outlined in this manuscript are freely available at https://github.com/ShixiangWang/prad_signature. Analyses can be read online at https://shixiangwang.github.io/prad_signature/.

## Acknowledgement

We thank Raymond Shuter for editing the text. Thank ShanghaiTech University High Performance Computing Public Service Platform for computing services. Thanks also to other members of Liu lab for helpful discussion.

## Author Contributions

SW collected the data, performed the programming and statistical analysis; HL, MS participated in data collection, preprocessing, and discussion; ZH, TW, XW, ZT, KW participated in critical project discussions; SW and XSL analyzed and interpreted the data; XSL supervised the study and wrote the manuscript.

## Supplemental Figure legends

**Figure S1.**
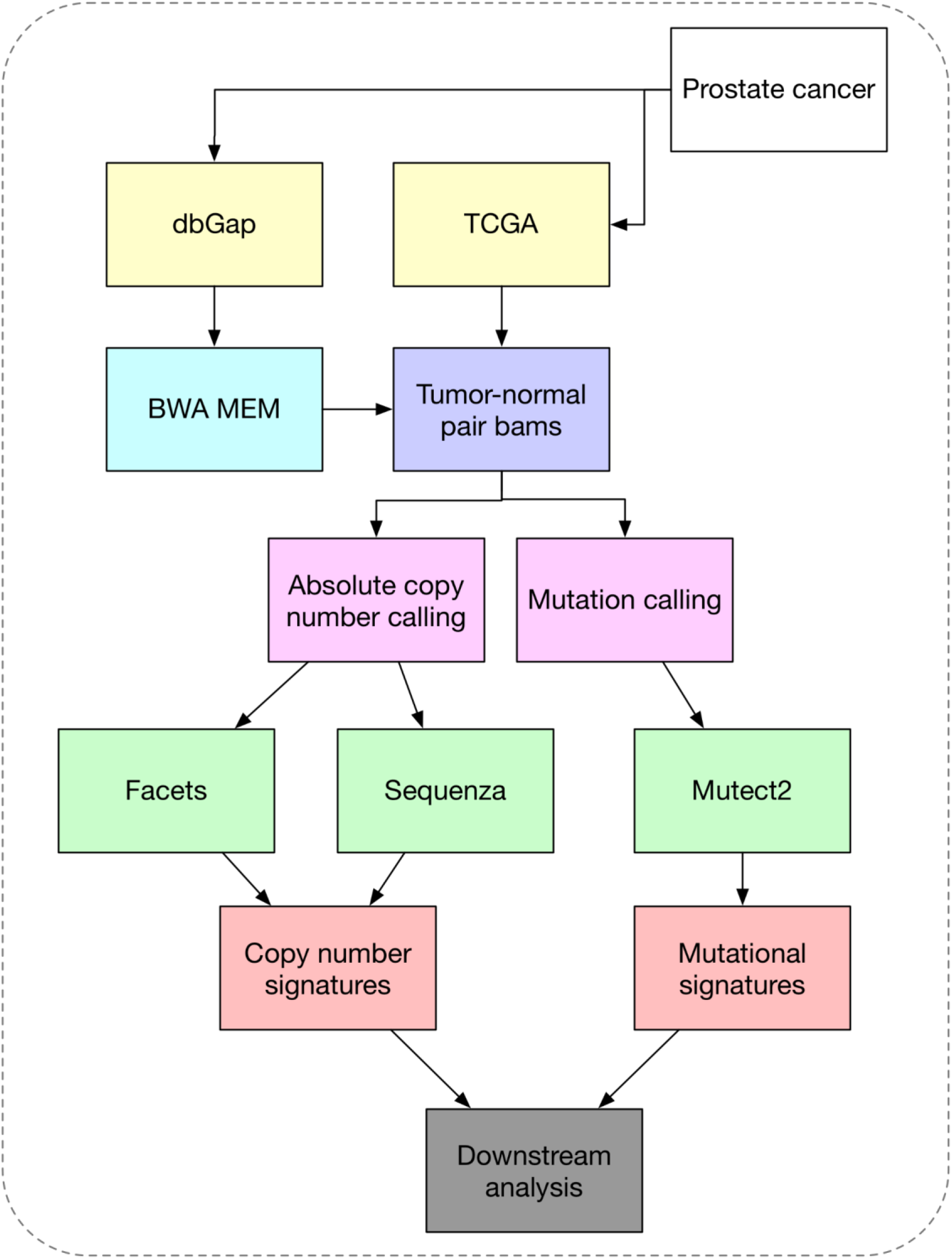
**Study design and flowchart depicting the basic processing steps from raw sequencing data to genome alteration signatures**.

**Figure S2.**
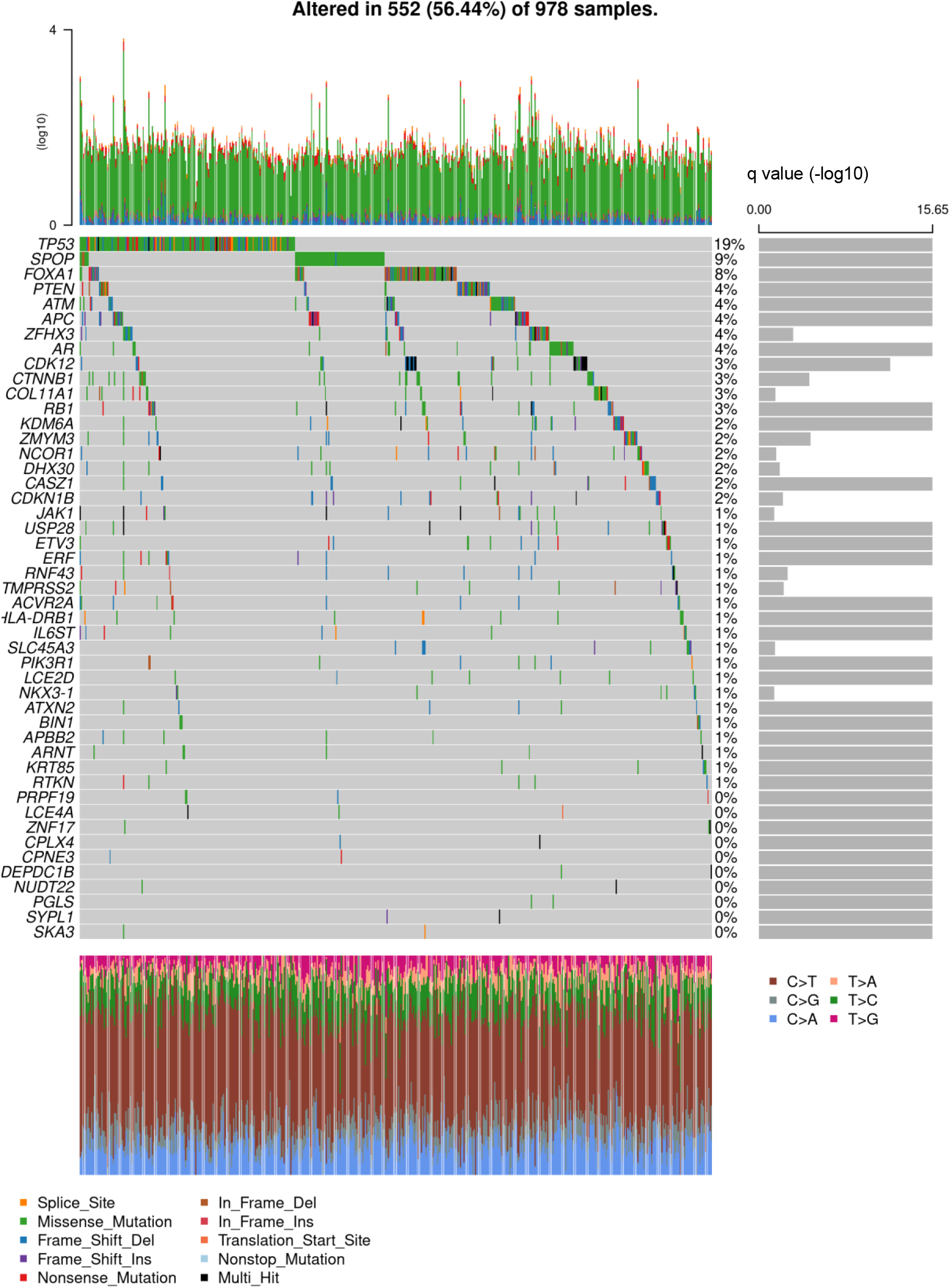
Mutational landscape of PC WES datasets. Driver genes are identified by MutSig with q value < 0.05. The right panel indicates log10 based MutSig q values for driver genes. This plot was generated by Maftools with default setting.

**Figure S3.**
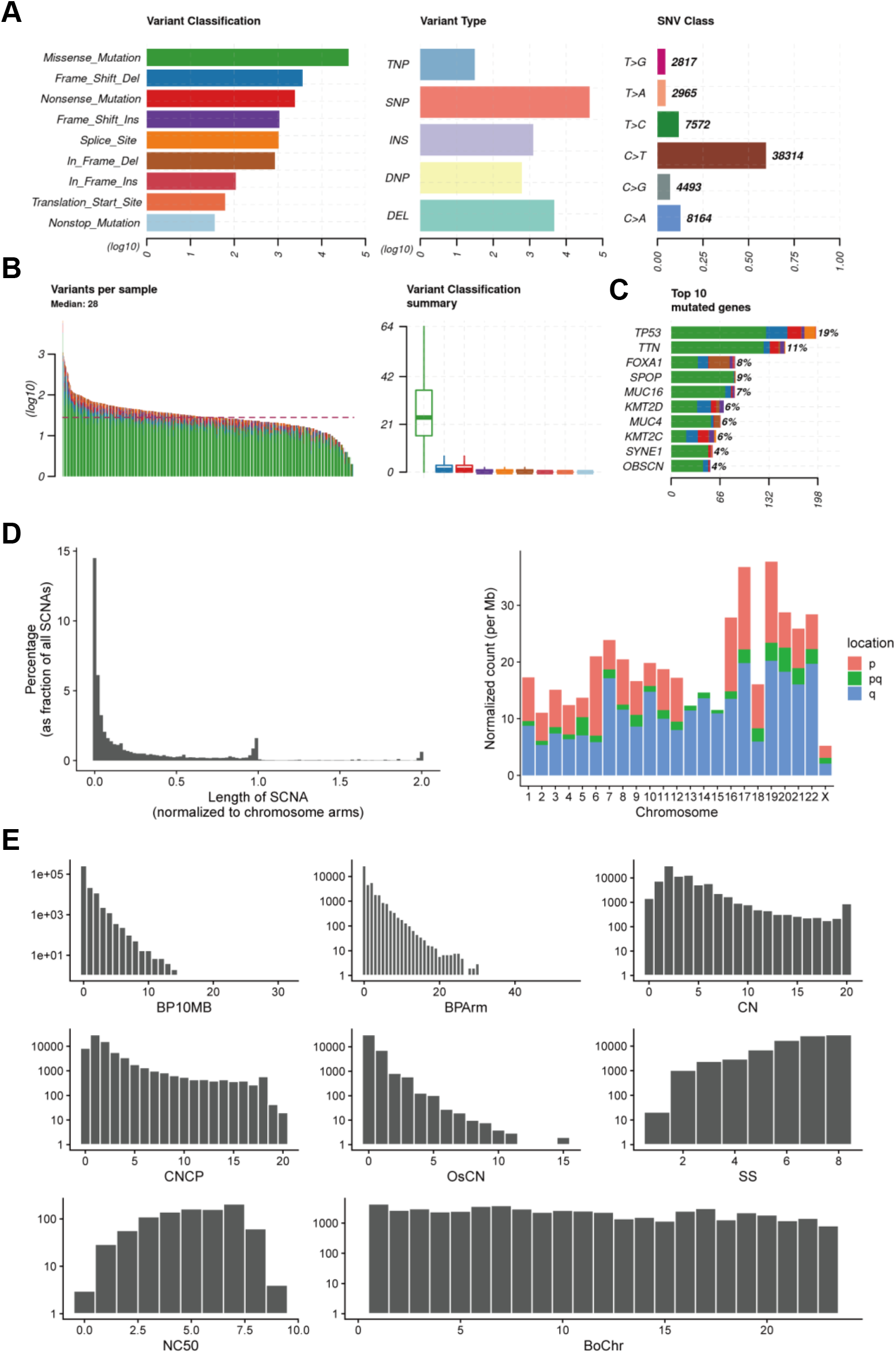
Summary for small scale variants and copy number alteration features in PC WES dataset. (A) Number of variants of different types in PC WES datasets. (B) Number of variants in each sample as a stacked barplot and variant classification as a boxplot. (C) Top 10 mutated genes as a stacked barplot by variant classification. (D) Length distribution of somatic copy number alterations (SCNA) segment and chromosome distribution of SCNAs. Location ‘pq’ represents a segment across both p arm and q arm. (E) Frequency distribution of 8 copy number features. The × coordinate 23 in feature ‘BoChr’ represents chromosome X.

**Figure S4.**
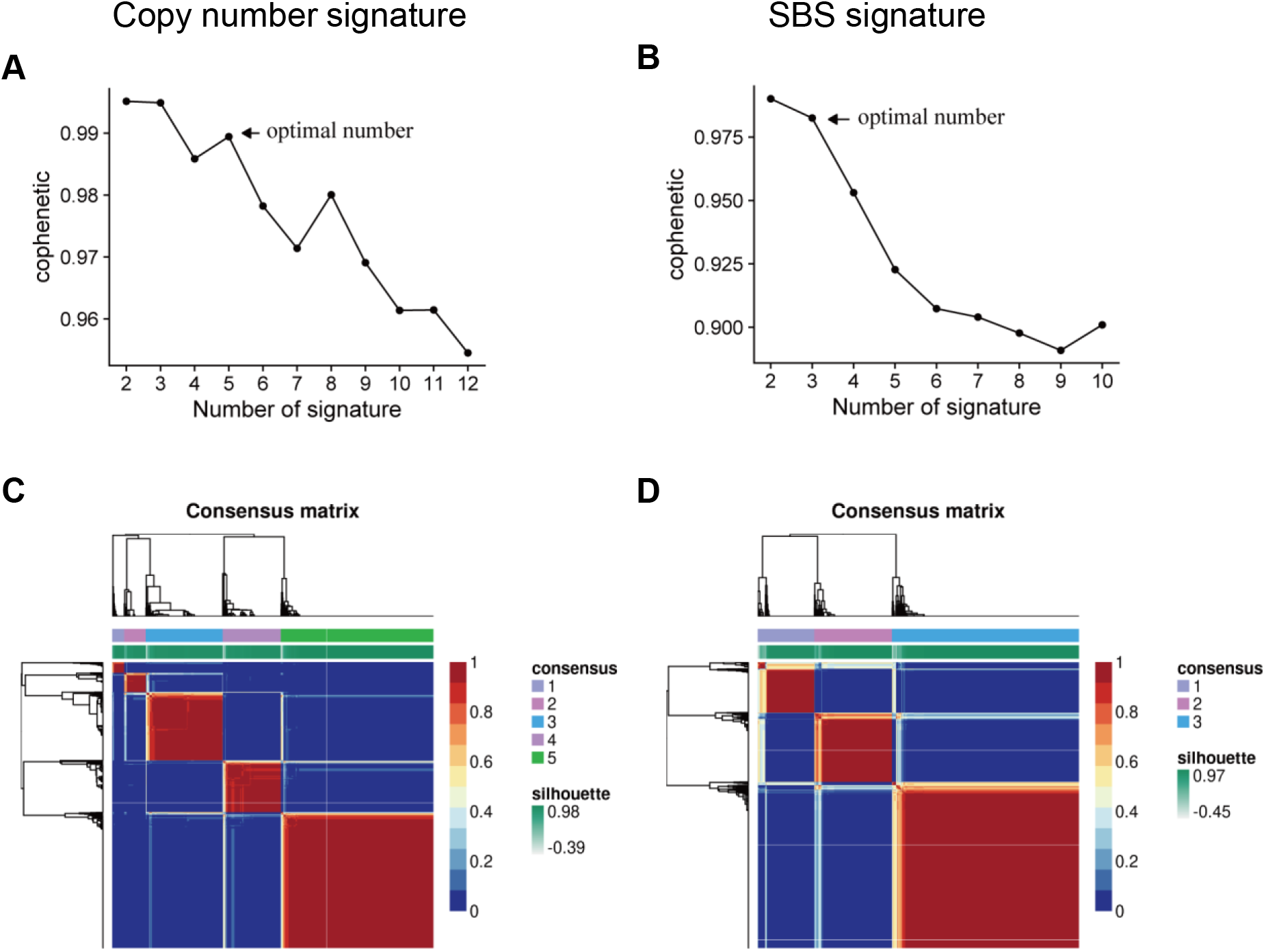
Cophenetic plot for determining the number of signatures. (A, B) Cophenetic plot analysis for copy number signatures (A) and SBS mutational signatures (B). (C, D) The consensus matrices for copy number signatures (C) and SBS mutational signatures (D).

**Figure S5.**
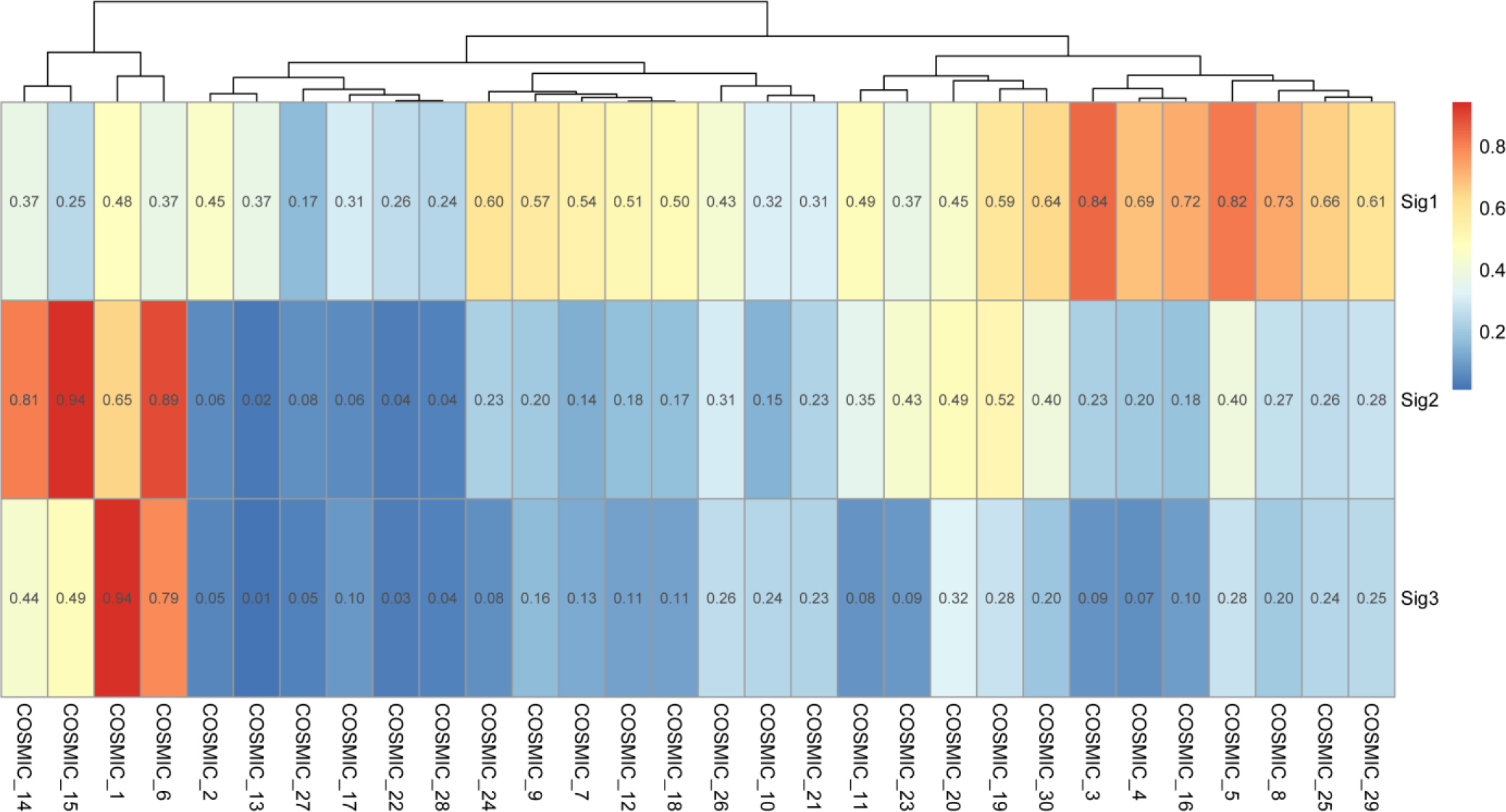
Cosine similarity analysis between SBS signatures identified in this study and COSMIC signatures.

**Figure S6.**
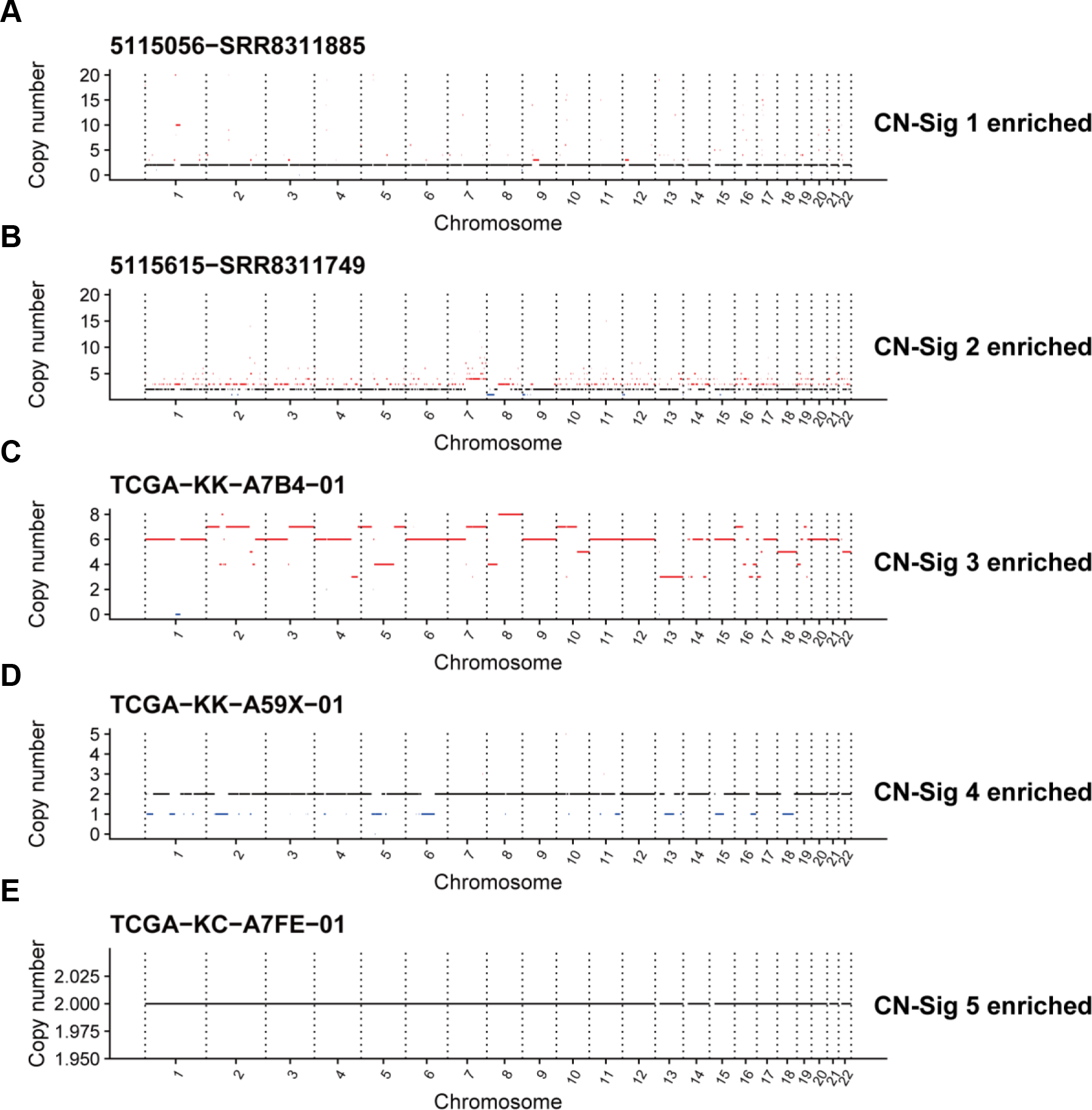
Representative absolute copy number profile for each copy number signature. Selected samples with enriched copy number signature CN-Sig 1 (A), CN-Sig 2 (B), CN-Sig 3 (C), CN-Sig 4 (D) and CN-Sig 5 (E) are shown. The segments with copy number gain and loss are labeled by red and blue color, respectively.

**Figure S7.**
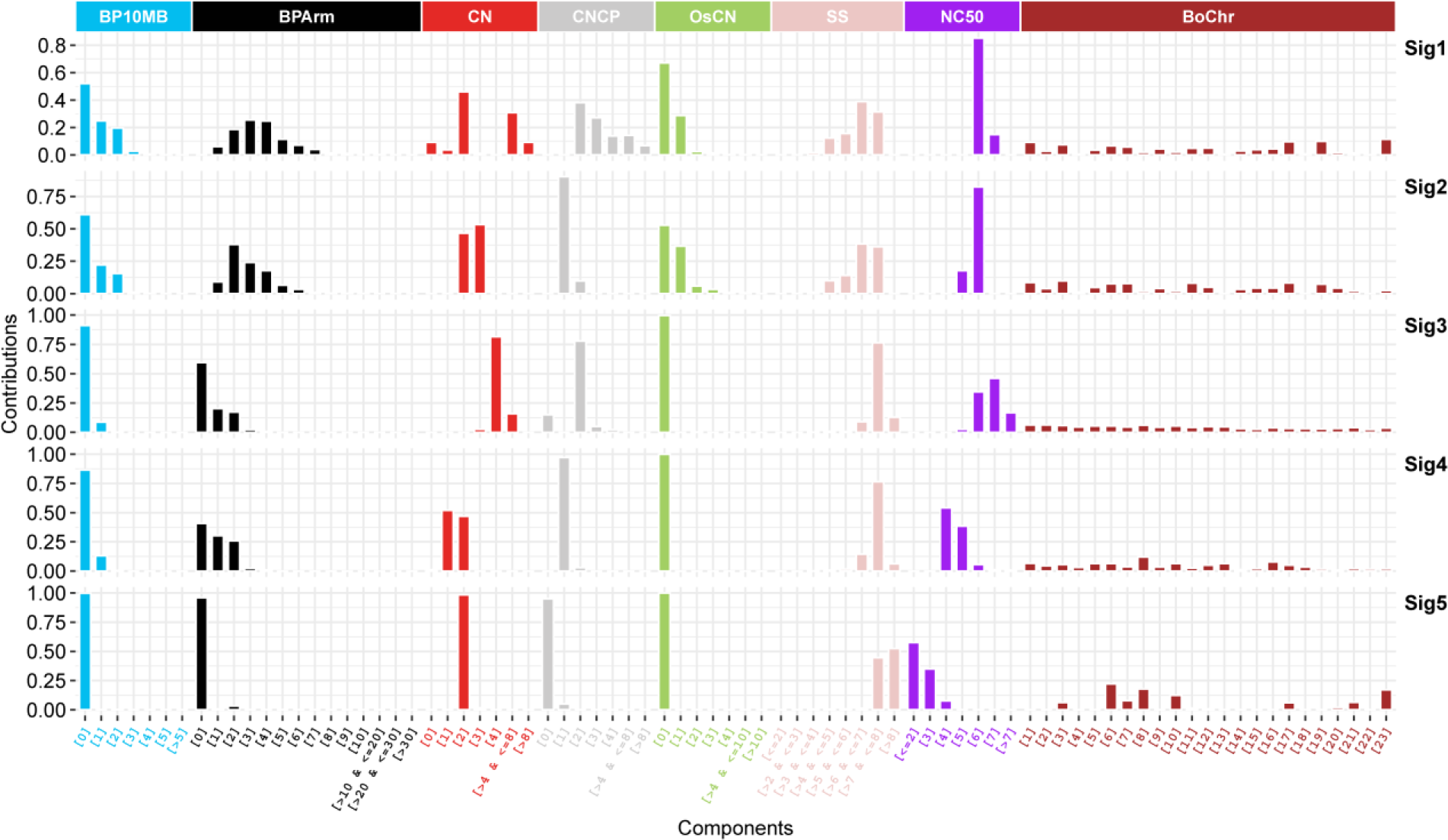
Copy number signatures identified in an independent MSKCC2020 prostate cancer dataset (n=1302).

**Figure S8.**
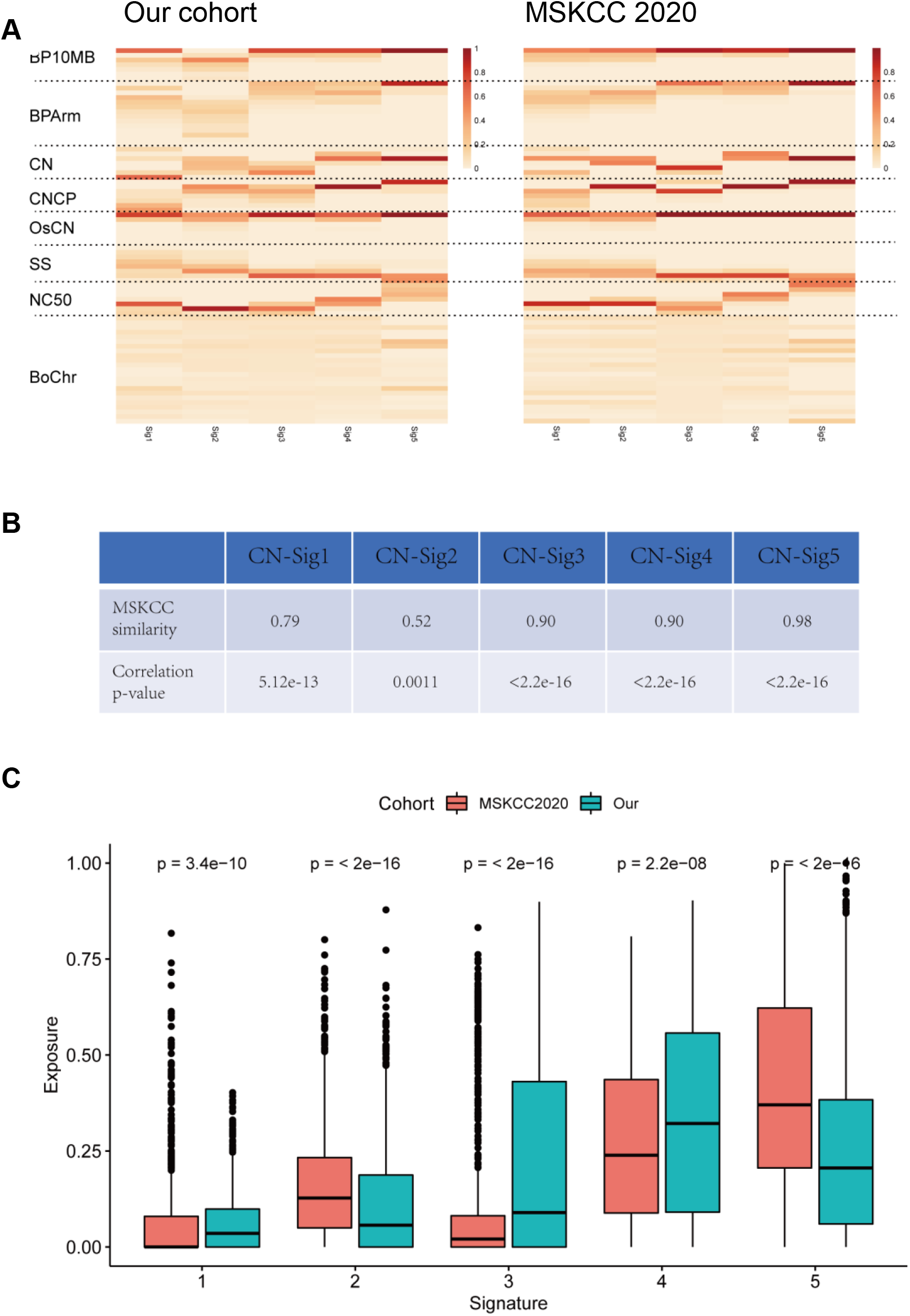
Validation of copy number signatures in an independent MSKCC2020 PC dataset. (A) Heat maps show component weights for copy number signatures in our cohort and an independent MSKCC2020 cohort. (B) Cosine similarity analysis of the copy number signature between our cohort and MSKCC2020 cohort. (C) Comparison of copy number signature exposures in our cohort and MSKCC2020 cohort. Significant differences are highlighted using asterisks (*P < 0.05, **P < 0.01, ***P < 0.001).

**Figure S9.**
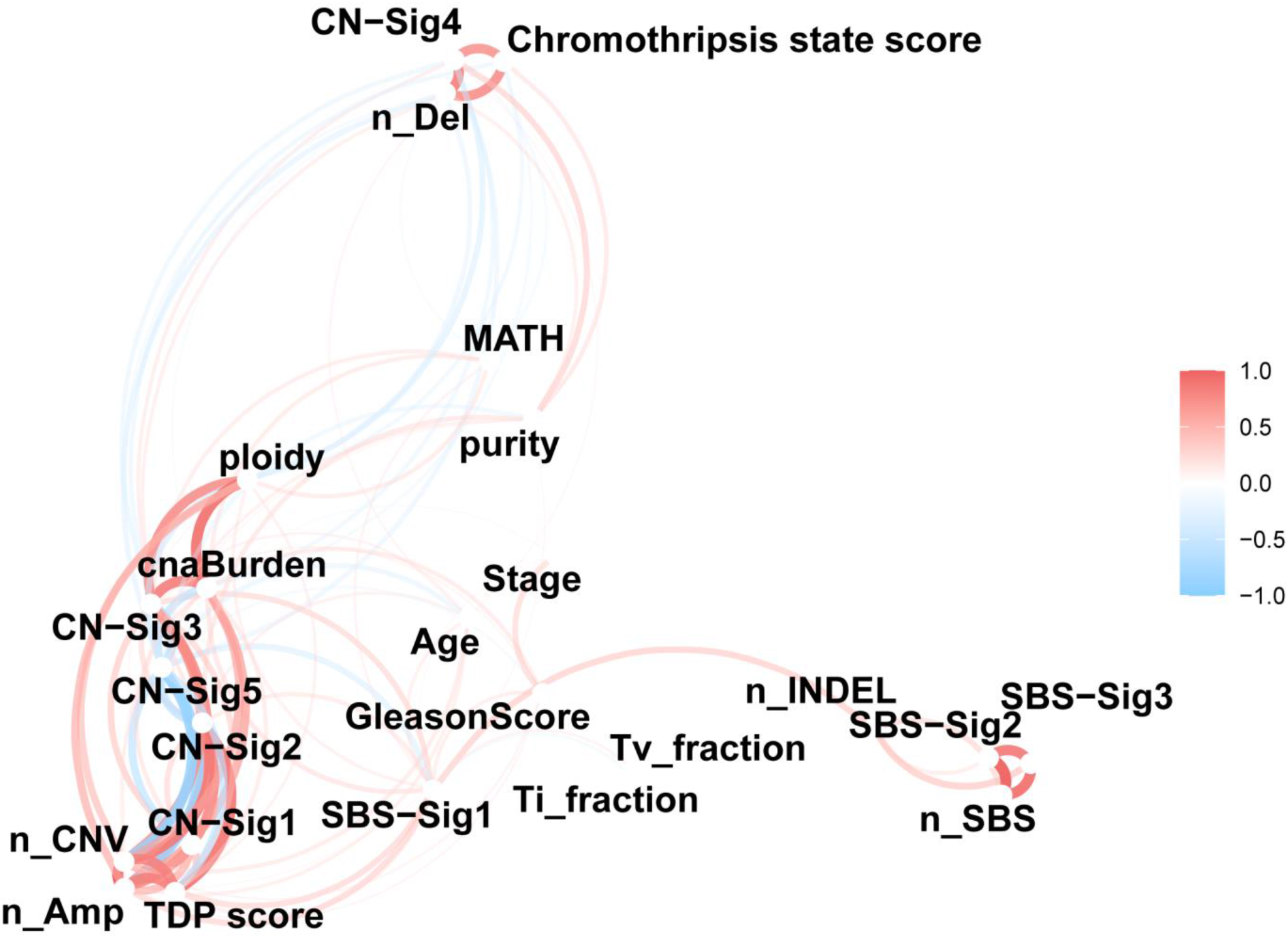
Correlation network analysis for genome alteration signatures. Variables that are more highly correlated appear closer together and are joined by stronger curves. Red color indicates positive correlation and blue color indicate negative correlation. The proximity of the points are determined using multidimensional clustering. Associations with Pearson correlation coefficient r>0.2 are shown.

**Figure S10.**
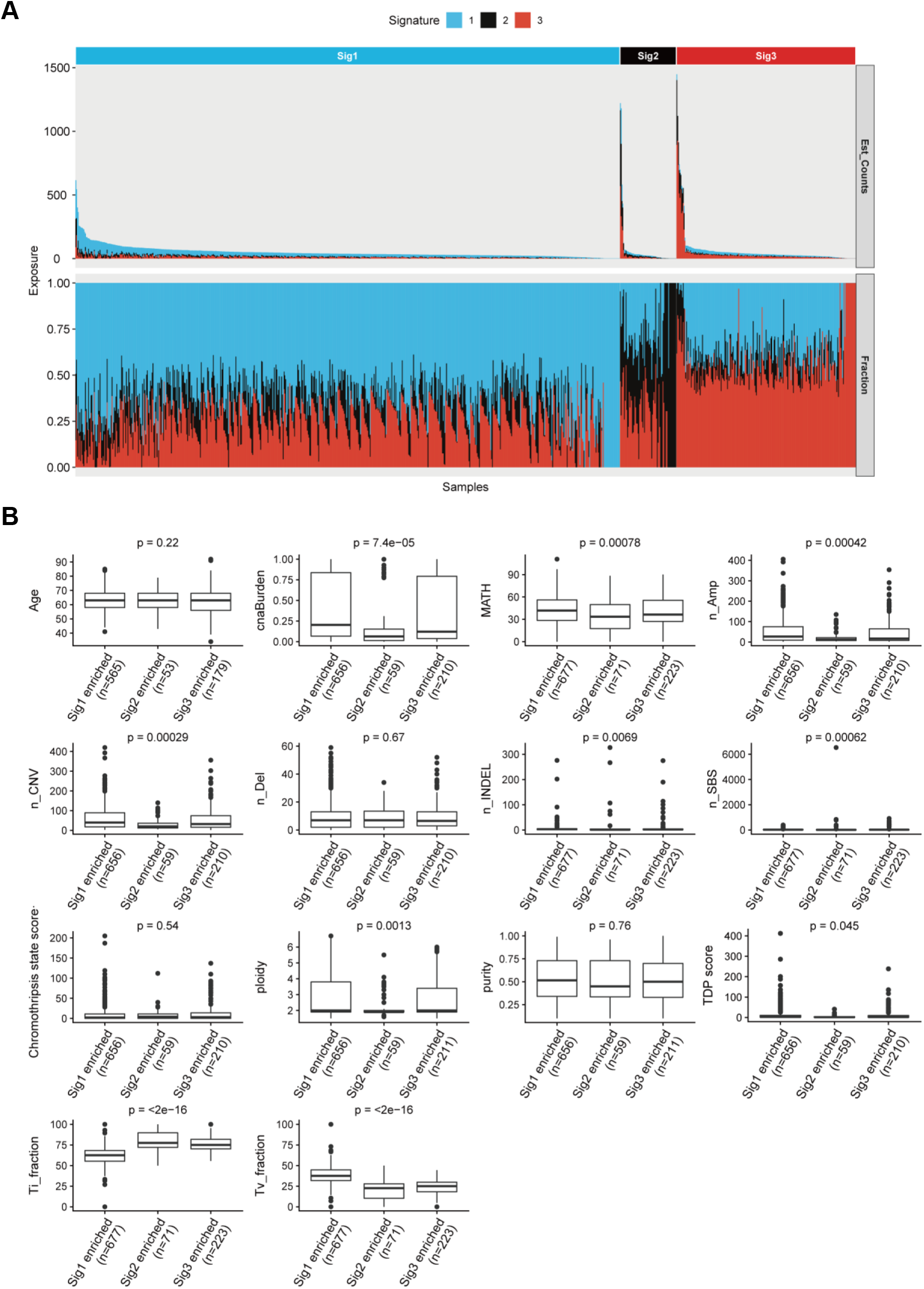
Sample clustering and heterogeneity analysis of PC based on the exposures of SBS signatures. (A) For each PC patient, the relative contribution (bottom panel) and estimated SBS mutational counts (top panel) of each signature are shown as a staked barplot. PC samples are clustered into three groups based on the consensus matrix from multiple NMF runs, and each group is specified by one enriched SBS signature. (B) Quantification comparison for somatic and clinical parameters among each SBS signature enriched PC group by boxplot. ANOVA p values are shown. Abbr.: n_, number of, e.g. n_SNV, number of SNV; INDELs, small insertions and deletions; Ti_fraction, transition fraction; Tv_fraction, transversion fraction; Amp, copy number segments with amplification; Del, copy number segments with deletion; TDP score, tandem duplication phenotype score; cnaBurden, copy number alteration burden; MATH, MATH score is a quantitative measure of intra-tumor heterogeneity.

**Figure S11.**
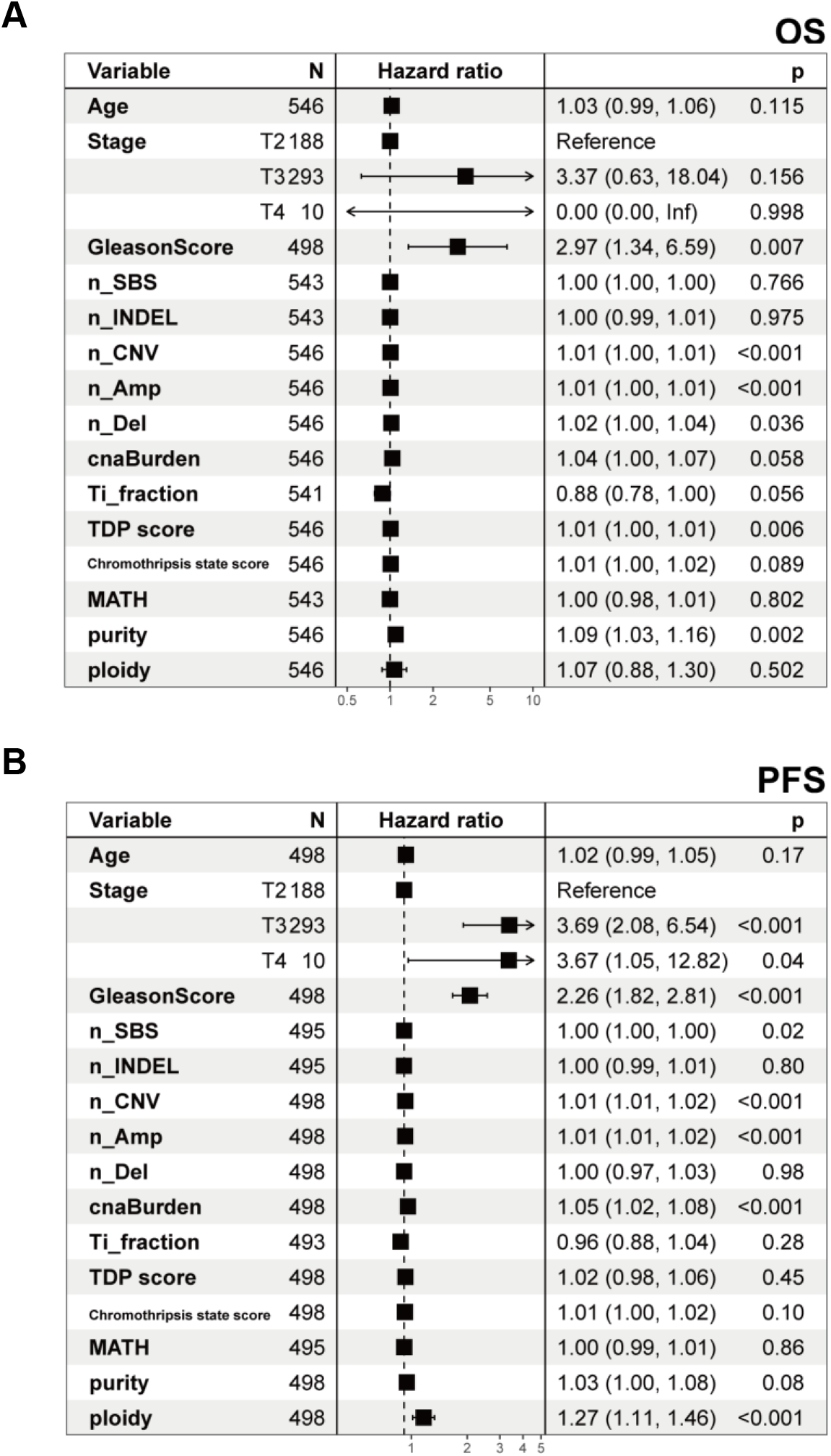
Genomic and clinical features and PC survival. Forest plots showing the relative risk of selected genomic and clinical features in overall survival (OS) (A) and progression-free survival (PFS) (B). Hazard ratios and p values were calculated by univariable Cox analysis. Abbr.: n_, number of, e.g. n_SNV, number of SNV; INDELs, small insertions and deletions; Ti_fraction, transition fraction; Tv_fraction, transversion fraction; Amp, copy number segments with amplification; Del copy number segments with deletion; TDP score, tandem duplication phenotype score; MATH, MATH score is a quantitative measure of intra-tumor heterogeneity.

**Table S1. Copy number component parameter setting**.

**Table S2. Cohort characteristics**.

**Table S3. Summary statistics for somatic genome variations**.

**Table S4. Selected pathways and genes for association analysis**.

**Table S5. Sequencing depth information for BAM files**.

**Table S6. Summary table for absolute copy number profiles detected in this study**.

**Table S7. Summary table for somatic mutations detected in this study**.

